# Developing an interactive, personalized patient decision aid for COVID-19 vaccination in Canada: Insights from a human-centered design and development study

**DOI:** 10.1101/2025.10.07.25337102

**Authors:** Doriane Etienne, Patrick Archambault, Isaac Bogoch, Christine T. Chambers, Andrea D. Chittle, Juliette Demers, S. Michelle Driedger, Eve Dubé, Marie-Pierre Gagnon, Teresa Gavaruzzi, Anik Giguere, Nathalie Grandvaux, Kelly Grindrod, Hina Hakim, Samira Jeimy, Jason Kindrachuk, Annie Leblanc, Shannon E. MacDonald, Ruth Ndjaboue, Magniol Noubi, Rita Orji, Jean-Sébastien Paquette, Elizabeth Parent, Jean-Sébastien Renaud, Beate Sander, Monica Taljaard, Dana Tannenbaum Greenberg, Marie-Claude Tremblay, Sabina Vohra-Miller, Vivian Welch, Holly O. Witteman

**Affiliations:** Department of Family and Emergency Medicine, Faculty of Medicine, Laval University; Vitam Research Centre for Sustainable Health; Research Centre of the CHU de Québec; Department of Family and Emergency Medicine, Faculty of Medicine, Laval University; Vitam Research Centre for Sustainable Health; Centre de recherche intégrée pour un système apprenant en santé et services sociaux du Centre int & eac; Divisions of General Internal Medicine & Infectious Diseases, Toronto General Hospital, University Health Network; Departments of Psychology and Neuroscience & Pediatrics, Faculties of Science and Medicine, Dalhousie University; Centre for Pediatric Pain Research, IWK Health; Department of Family Medicine, McMaster University; Department of Biochemistry, Faculty of Science, McGill University; College of Community and Global Health, Rady Faculty of Health Sciences, University of Manitoba; Research Centre of the CHU de Québec, Laval University; Faculty of Nursing Sciences, Laval University; Vitam Research Centre for Sustainable Health; Department of Medical and Surgical Sciences, University of Bologna; Vitam Research Centre for Sustainable Health; Department of Family and Emergency Medicine, Faculty of Medicine, Laval University; Department of Biochemistry and Molecular Medicine, Faculty of Medicine, Université de Montréal; Centre de recherche du Centre hospitalier de l’Université de Montréal (CRCHUM); School of Pharmacy, Faculty of Science, University of Waterloo; Department of Family and Emergency Medicine, Laval University; VITAM Research Centre for Sustainable Health; Research Centre of the CHU de Québec; Division of Clinical Immunology and Allergy, Dept of Medicine, Western University; Department of Medical Microbiology & Infectious Diseases, Rady Faculty of Health Sciences, University of Manitoba; Department of Family and Emergency Medicine, Faculty of Medicine, Laval University; Vitam Research Centre for Sustainable Health; Faculty of Nursing, University of Alberta; School of Social Work, Faculty of Letter and Human Sciences, Université de Sherbrooke; Research Centre on Ageing; Faculty of Computer Science, Dalhousie University; Department of Family and Emergency Medicine, Faculty of Medicine, Laval University; Department of family medicine and emergency medicine, Faculty of Medicine, Laval University; Vitam Research Centre for Sustainable Health; Health Systems and Policy Research Collaborative Centre, University Health Network; Methodological and Implementation Research, Ottawa Hospital Research Institute; Diabetes Action Canada; Department of family medicine and emergency medicine, Faculty of Medicine, Laval University; Dalla Lana School of Public Health, Division of Clinical Public Health; Bruyere Health Research Institute

**Keywords:** COVID-19 vaccination, patient decision aid, web-based tool, iterative human-centred design, digital health, health literacy, user engagement, public health communication, data visualization, personalized avatar

## Abstract

**Background:** The COVID-19 pandemic highlighted the need for practical digital health tools to support informed decision-making amidst rapidly evolving evidence and widespread misinformation.

**Objective:** We iteratively developed and refined VaxDA-C19, a bilingual (English and French) web-based patient decision aid designed to support informed decision-making in Canada about COVID-19 vaccination. VaxDA-C19 integrates interactive and personalized features aimed to enhance vaccine confidence, reduce cognitive overload, and respond to diverse informational needs.

**Methods:** We developed VaxDA-C19 using an iterative, user-centered design approach. Throughout the development process, we involved a citizen panel, healthcare professionals, user experience designers, and scientific experts to guide refinements. We also conducted usability testing sessions with adults in Canada, using semi-structured interviews, comparative testing, and think-aloud protocols with thematic analysis. We ultimately conducted four design cycles in total: three with adults in Canada (cycle 1: n=9 users; cycle 2: n=22 users; cycle 3: n=3 users), one overlapping and one additional cycle with expert reviewers (cycle 3: n=5; cycle 4: n=9).

**Results:** In Cycle 1, user feedback guided design decisions about how to present quantitative information and technical vaccine descriptions more simply. In Cycle 2, while most users (82%) favored in-depth explanations of vaccine development, a few raised concerns about content that could be perceived as politically charged. Cycle 3 identified usability improvements, including more explicit navigation controls, simplified medical terminology, and optimized interactive components (avatars, sliders). Expert reviews in Cycle 4 refined linguistic consistency, mobile responsiveness, content transparency, and scientific accuracy, emphasizing explicit instructional guidance and bilingual accessibility.

**Conclusions:** Our iterative process produced a personalized, bilingual digital decision aid to support evidence-informed, values-congruent decisions about COVID-19 vaccination. A randomized controlled trial will further evaluate VaxDA-C19’s impact on vaccination intentions, knowledge retention, emotional responses, decisional conflict, and decisional regret. If it proves effective, the patient decision aid may also be used as a platform to support other vaccine decisions, namely, influenza, measles, shingles, pertussis, and potentially other emerging infectious diseases.

## Introduction

The COVID-19 pandemic exposed challenges in public health decision-making driven by rapidly-evolving scientific evidence and widespread misinformation [1–8]. People had to make complex choices, such as whether to accept vaccination or adopt preventive behaviours, often under conditions of uncertainty [4,9–13]. These challenges highlighted the need for decision support tools that empower users to make values-congruent choices informed by the best available evidence [1,10,14–21].

Patient decision aids support shared decision-making by making decisions explicit, providing balanced and evidence-based information about options, and helping individuals clarify their values [22]. Robust evidence from meta-analyses and systematic reviews, including a comprehensive Cochrane review, demonstrates that patient decision aids significantly improve knowledge, accuracy of risk perceptions, and alignment between decisions and personal values, while also reducing decisional conflict [23,24]. In the context of vaccination, a systematic review and meta-analysis of five randomized controlled trials (total N=2158) found that patient decision aids for vaccine decisions had significant positive effects on both vaccine intentions (OR 1.89, 95% CI: 1.20–2.97) and uptake (OR 1.77, 95% CI: 1.25–2.52) [25].

Emerging evidence suggests that interactive and personalized digital tools can encourage behaviour change among individuals who have not yet engaged in recommended practices, reinforce existing behaviours, and provide reassurance [26–29]. Such multiple purposes may be particularly critical during periods of crisis, when individuals seek validation for their actions and guidance for future decisions [30,31].

As COVID-19 vaccines move towards regular annual or biannual vaccines in Canada, similar to influenza vaccines, there is a need for patient decision aids that can be easily updated to reflect updated vaccines and recommendations. To this end, we created VaxDA-C19, a web-based vaccine (Vax) patient decision aid (DA) designed to support COVID-19 (-C19) vaccination decisions in Canada. VaxDA-C19 is designed to support multiple languages and to provide information in a layered way, meaning that people who want to quickly access basic information can view brief bullet points while those who want to know more can expand sections, get more details, and access original references. Our goals were to provide clear, evidence-based information on vaccination options, benefits, and risks to oneself and others while reducing cognitive overload. The present paper describes VaxDA-C19’s design and development process.

## Methods

### Overview and theoretical frameworks

Our overall design process followed the International Patient Decision Aids Standards [32]. The patient decision aid was structured according to the Ottawa Decision Support Framework [33]. The web-based design and development approach combined human-centered design principles [34] and a Scrum framework [35]. Human-centered design prioritizes direct user involvement from the early stages and continuously adapting development based on user feedback [34]. Scrum employs short, iterative “sprints” or short development cycles in which teams hold short, frequent meetings to coordinate progress and align development priorities with emerging user feedback.

### Citizen panel

To help guide the overall project, a citizen panel (n=6), comprising people from a range of age groups, languages, educational backgrounds, health statuses, and geographic regions (Alberta, Saskatchewan, Ontario, Québec) provided regular feedback about the design and development of the patient decision aid and other interventions (reported separately) as well as the user testing methods and results. In the context of COVID-19 restrictions in place at the beginning of the project, the citizen panel was assembled from existing contacts with patient and citizen partners across a range of prior projects and life circumstances. This group met periodically by Zoom for two hours at a time to discuss ongoing project design, development, and study planning. This group served as an advisory panel, distinct from user testing with previously unknown members of the general public, described below [34].

### Expert involvement

Healthcare professionals and research experts, including infectious disease researchers, epidemiologists, primary care providers, and two user experience designers (internal and external experts), participated in the iterative process as expert reviewers. Depending on their exact area of expertise, their roles were to help ensure the accuracy of the patient decision aid content and to review the user experience of the web-based patient decision aid [34].

### Ethics

This study received ethical approval from the “Comité d’éthique de la recherche en sciences de la santé” of the Université Laval (approval no: 2022-498 A-1 and 13-01-2025 and 2020-222). All participants received detailed study information and provided written informed consent before participation.

### Recruitment for user testing

In light of COVID-19 restrictions in place during the bulk of this project, we used online tools to conduct our user tests. Specifically, we recruited user testing participants through tailored Facebook advertisements, then conducted user testing sessions using Zoom, with participants sharing their screen with the interviewer. Eligible participants for user testing were adults (≥18 years) residing in Canada, able to communicate in English or French, and possessing basic computer skills. Individuals without internet access or unable to provide informed consent were excluded. We therefore selected Facebook options to advertise to people who were 18 years and over across Canada, and who indicated an education level of either associate degree, high school graduate, or unspecified. We did not exclude people who had other educational statuses, but used tailored recruitment in an effort to include people across a range of backgrounds, as our previous experiences suggested that people who respond to such advertisements are often more highly educated. We stopped recruiting for each cycle after reaching thematic saturation in comments and usability issues identified. We offered user testing participants CAD $40 (US$28.80) for their time and to help defray internet costs.

### Overall design of the intervention

We used Figma design software to iteratively design the overall structure and appearance of VaxDA-C19. As described further in Results, the patient decision aid ultimately included an introductory section in which the patient decision aid is presented and the user selects the age group of the person for whom a decision is required (themself or their child), their province or territory of residence (which determines which vaccine(s) are available), and the decision they are making (whether or not to get a primary series of vaccines, whether or not to get a booster dose, which vaccine to get, or which booster to get.) The user is then invited to create an avatar representing themself or their child. This avatar is used in the next section, which describes the individual-level benefits and risks of the vaccine. The top of the page provides a summary, and the user can click to view more detailed explanations below the summary. On the next page, the user can choose to create up to 8 more avatars representing people around them or their child or to have additional avatars auto-generated. They may then view or skip a brief (2.5-minute) animation showing how vaccination may protect those around us [36]. Following this optional component, the user is invited to reflect on what matters to them, relevant to the decision, using a values clarification method previously developed and tested in the context of treatment decisions and influenza vaccination decisions [37]. Finally, the user has the option to read answers to frequently asked questions and to learn more about our team.

### Design and development approach

Our multidisciplinary team followed a human-centred design over four iterative cycles, summarized in Figure 1 [34]. As per the International Patient Decision Aids Standards [32], our design process was structured in multiple testing cycles, each involving small to medium user samples (3–31 participants, no overlap between cycles). This aligns with cognitive ergonomics standards [38–40], which state that iterative testing with small samples efficiently identifies approximately 85% of usability issues per cycle. After each cycle, the team met to review findings and make decisions about modifications. The citizen panel met separately throughout the project to offer advice from their viewpoints.

**Figure 1.**
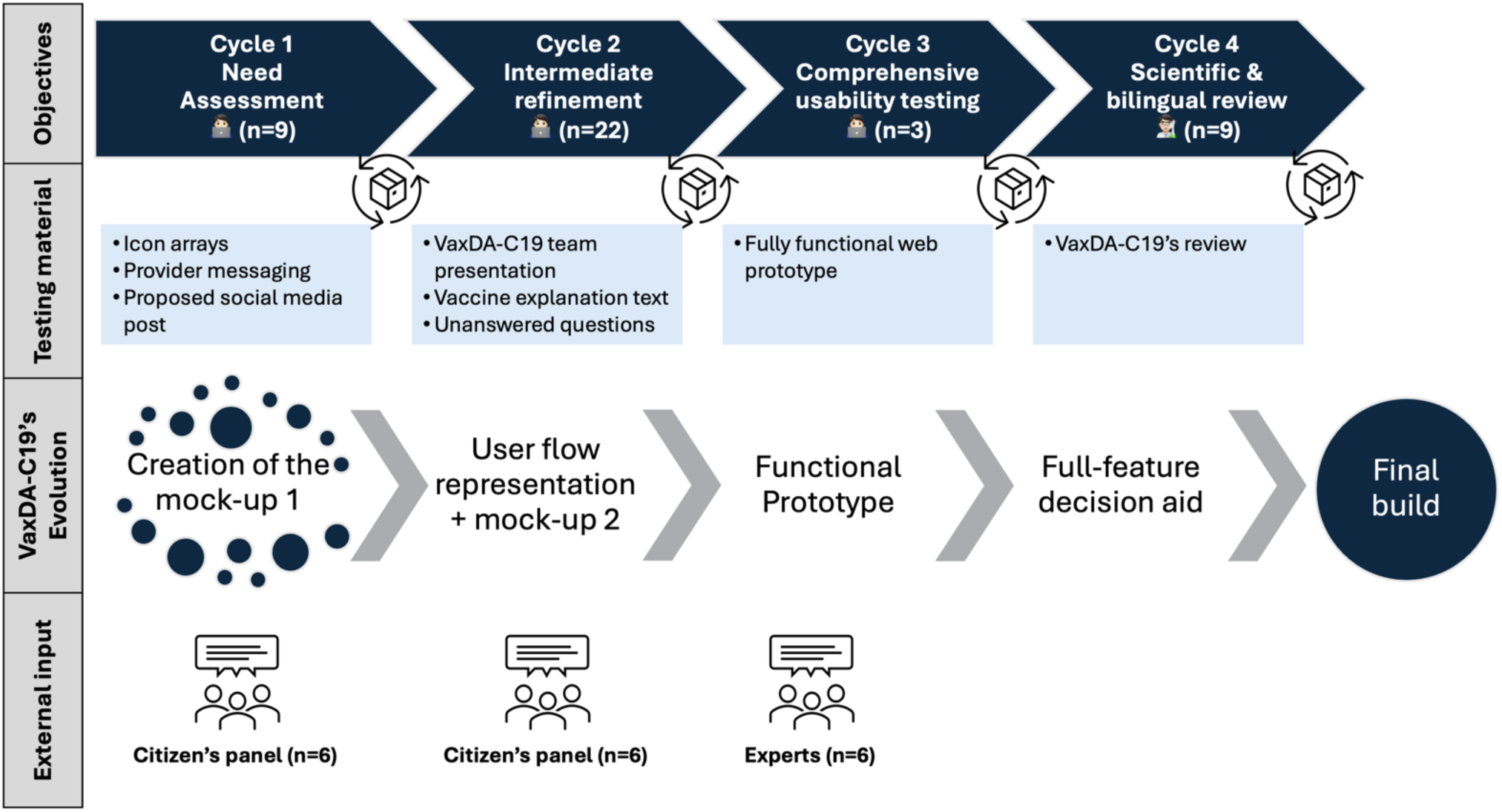
Iterative development cycles of VaxDA-C19, a web-based patient decision aid

### Data collection and analysis

We used Zoom to conduct user testing interviews. We evaluated whether VaxDA-C19 met its communication goals, then applied a rapid thematic analysis [41,42] to qualitative data from interviews, comparative testing and think-aloud sessions. Two trained research associates (EP, HH) independently coded transcripts, first extracting functional requirements (i.e., needed features, content adjustments, and navigation flows), and then flagging any usability issues. We resolved disagreements through discussion with the lead author (DE) and principal investigator (HW) until we reached a consensus. Finally, DE, EP, and HW organized the agreed functional requirements and identified bugs into a clear, detailed plan to guide the development team.

### Iterative Cycles

Across all cycles, we presented participants with increasingly higher-fidelity versions of VaxDA-C19 prototypes. We asked participants to summarize the information, identify points of confusion or unanswered questions, and react and respond to wording.

#### Cycle 1

For the needs assessment, we developed an initial content mock-up of VaxDA-C19 focused on primary COVID-19 vaccination (first-dose decision). We based the mock-up’s content on contemporaneous recommendations from the Canadian National Advisory Committee on Immunization, the Institut national de santé publique du Québec, and Health Canada authorizations. Feedback from the citizen panel and content experts addressed clarity, scientific accuracy, and relevance. We then recruited participants (n=9) via tailored Facebook ads and conducted individual semi-structured Zoom interviews, whose interview guides are shown in full in Appendices 1–2. In each session, we asked participants to interpret icon arrays, react to hypothetical provider messaging, and consider a social media post.

For the icon arrays, as shown in Figure 2, we offered two icon array styles for risk communication: avatars with colored shirts and avatars with circular backdrops, and asked participants to compare these, focusing on how well or poorly these visual aids conveyed risk data.

**Figure 2.**
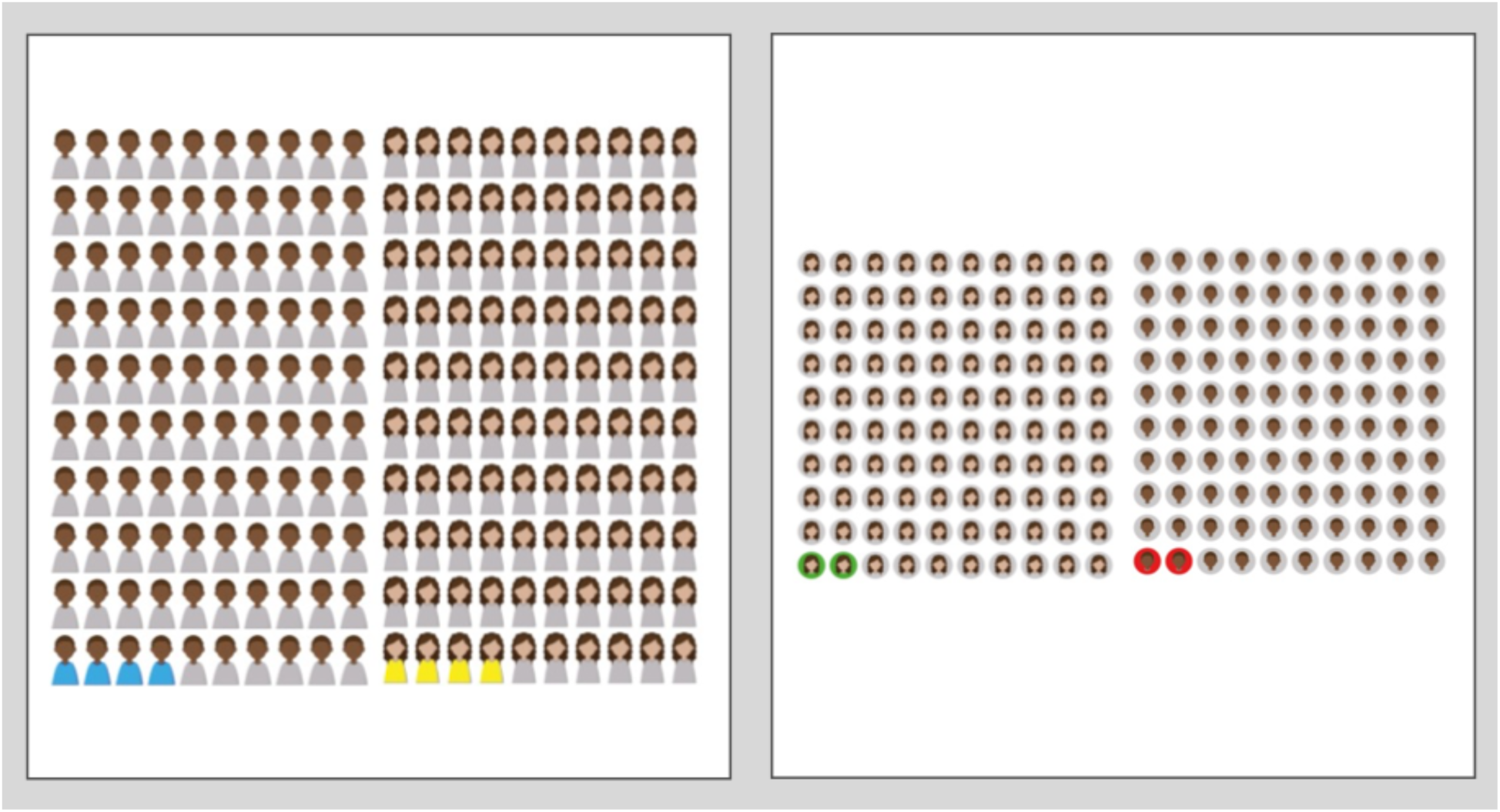
Icon arrays used during the first cycle

For the provider messaging, we asked people who were able to receive messages from their primary care provider, “Imagine your primary care health professional (e.g., family doctor, nurse practitioner) sends you a message like, ‘I got my COVID-19 vaccine when I was eligible. Now that you are eligible, I’d like to support you by answering any questions you might have. Here’s a link with some information: link. Please send me a message if you have more questions.’ How would that make you feel? How might you react or respond?” We asked participants who were unable to receive messages to respond to a similar message, but imagining it was during an in-person visit. Full details are shown in Appendix 1.

The proposed social media post aimed to explain why COVID-19 vaccines moved so rapidly through development, for use in a potential image-based post, such as an Instagram carousel. The proposed English text read: (1) 9 women can’t make a baby in 1 month, no matter how well they work together! (2) But when a bunch of the world’s best scientists work together, with enough research funding, motivated by a pandemic that is disrupting lives around the world, vaccine development can happen faster than usual. (3) Making a new vaccine usually involves months or years of waiting time. Waiting to get more research funding. Waiting for enough people to sign up for studies that make sure the vaccines are safe and work well. Waiting for approvals from independent regulatory authorities before starting to produce the vaccine for distribution. (4) Usually, this waiting happens because the need for the vaccine isn’t considered urgent enough for everyone to drop everything else and make the new vaccine everyone’s top priority. But COVID-19 has been a different story. It has been a priority around the world. (5) To make COVID-19 vaccines, they cut out a lot of the waiting. But they didn’t cut corners. All the safety checks are still being done. When COVID-19 vaccines are approved by Health Canada (Canada’s independent agency, staffed by top-notch scientists and not affiliated to any political party), you can trust that they are safe, and they work. (6) We are Canadian scientists, doctors, nurses, and experts in health and vaccines. None of us works for pharmaceutical companies. We don’t accept pharmaceutical funds either. We will get the COVID-19 vaccines for ourselves as soon as they are approved by Health Canada and available to us. Once they have been tested and approved for children, our children will get the vaccines, too. (7) Still have questions about COVID-19 vaccines and don’t know where to ask? We want to use our scientific training to help answer your questions. Send us your question here. We will do our very best to answer as many as we can. Full text is available in Appendix 2.

#### Cycle 2

Based on results from cycle 1, after identifying initial user needs, we developed the first version of content using Google Slides to create mock-ups of a proposed website. These mock-ups served as early prototypes, allowing us to conceptualize the user interface and gather feedback from the citizen panel on design, usability, and information presentation prior to investing substantial time in web development. Figure 3 maps the user flow. It began with a page of welcome text and instructions, then a page in which people indicate the age and province of residence of the person for whom a vaccine is being considered, then a page in which people build an avatar representing that person, then a page showing the individual-level risks and benefits of the vaccine for that person (using the avatar previously created to illustrate statistics), then a section showing the population-level benefits of vaccination, then a values clarification method to help people align their decision with what matters to them, and finally, frequently asked questions, information about where to obtain a vaccine, references supporting the content, and information about our team.

**Figure 3.**
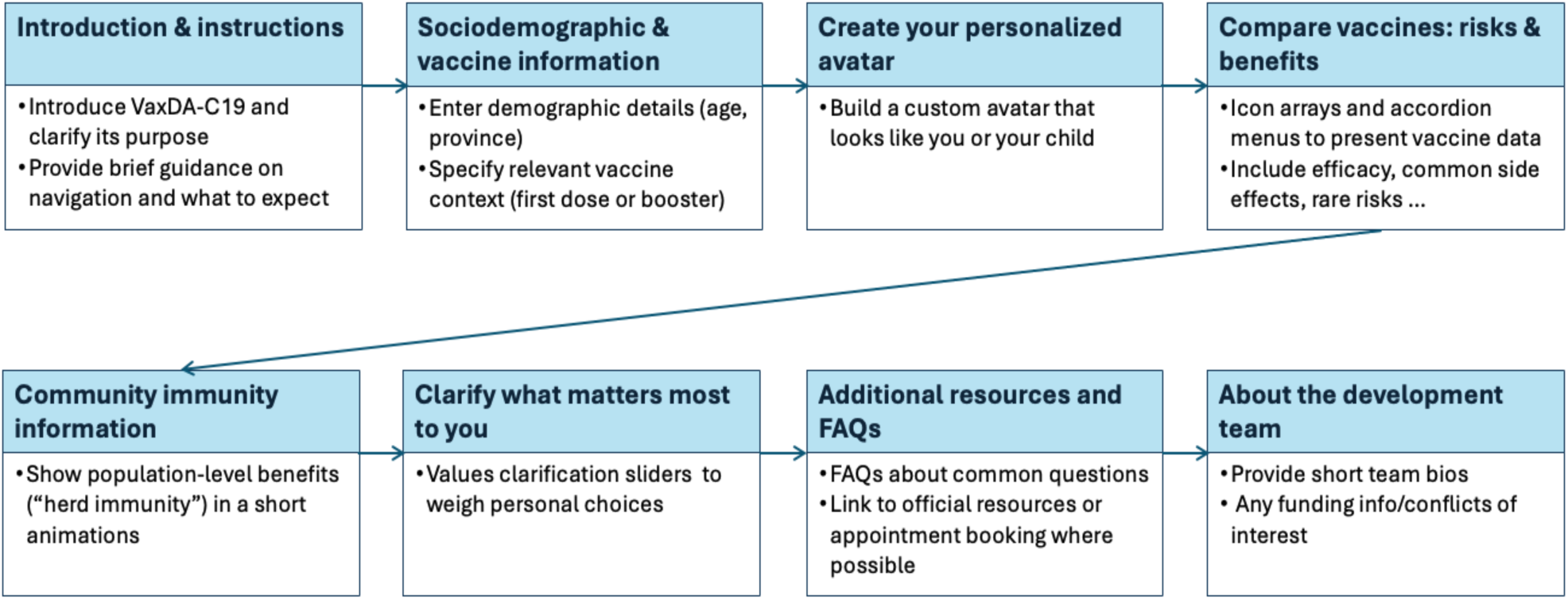
User flow representation of our web-based patient decision aid, VaxDA-C19

We then recruited new participants (n=22) via tailored Facebook ads and conducted a second round of individual semi-structured interviews on Zoom, again applying thematic analysis (Appendices 3–4). In each interview, we asked: “In your own words, what was this text about?”, “Why do you think these vaccines arrived so quickly?” and “What important questions about COVID-19 vaccines remain unanswered?” During these sessions, we used A/B testing to compare designs and content presentations of the team working on VaxDA-C19 and related projects. A/B testing, or split testing, involves presenting two versions (A and B) of a webpage or app to different user groups to determine which version performs better based on predefined metrics [43]. In this case, participants viewed two versions of the team presentation. Version A included only drawings, while Version B included photographs and the names of research team members. Figure 4 shows the two designs, and full details are available in Appendix 4.

**Figure 4.**
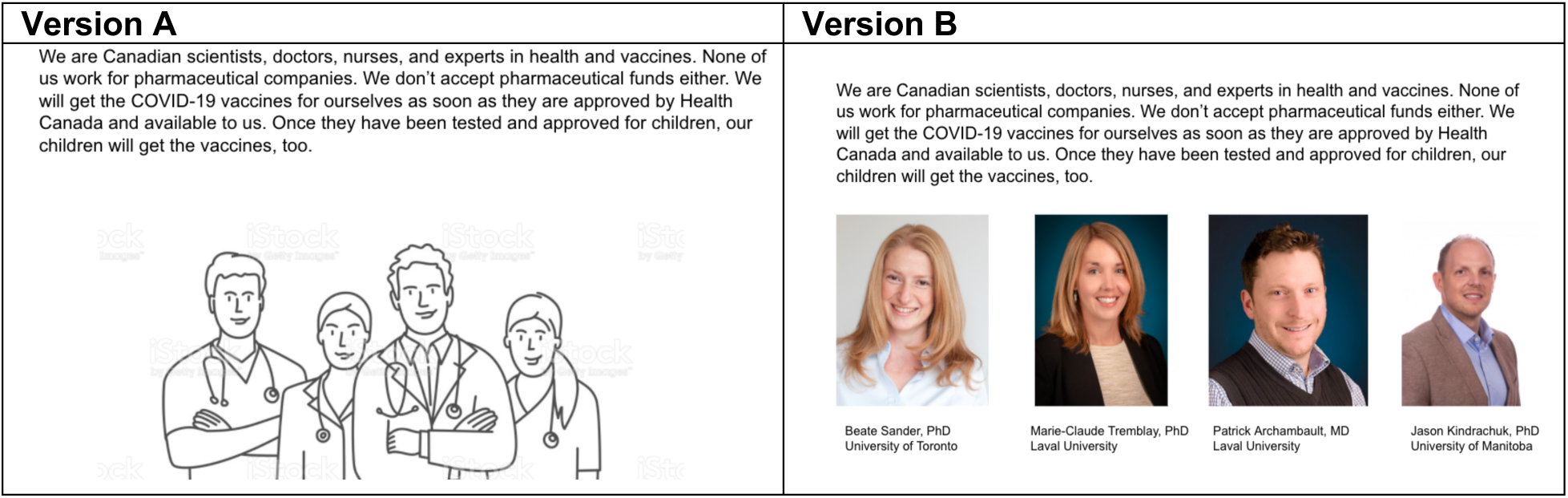
Comparative testing of user preferences for vaccine scientists’ presentation

**Figure 5.**
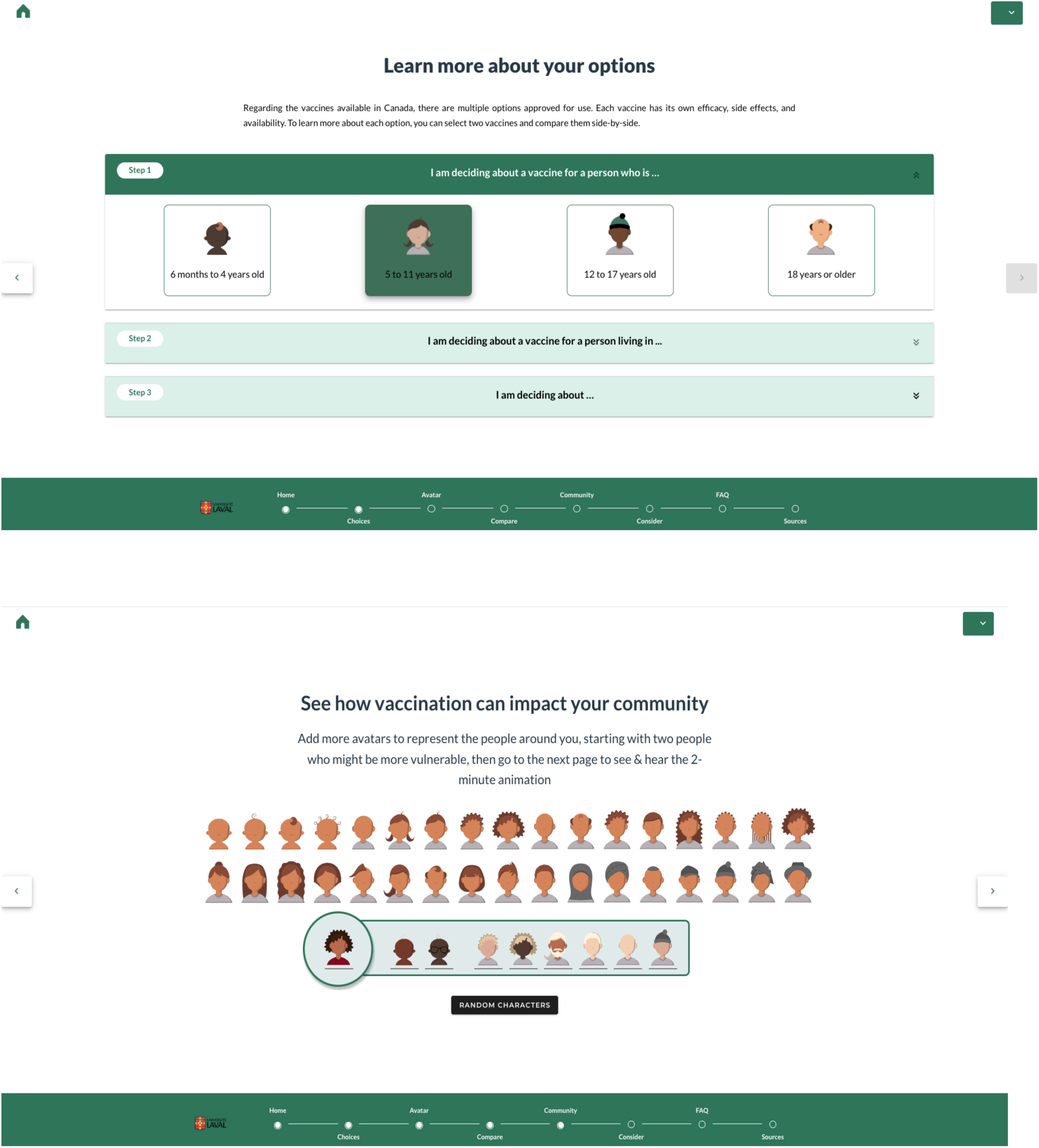
Selected views from the final version of VaxDA-C19

#### Cycle 3

Cycle 3 moved VaxDA-C19 from static mock-ups into a fully functional web prototype, then tested it with end users and experts. Based on results from cycle 2, we added five interactive features. These included having users create a personalized avatar to promote personalized user engagement [44,45] and to take advantage of the Proteus effect, a documented effect in which people identify with their avatar [46,47]. VaxDA-C19 then uses that personalized avatar in icon arrays to facilitate visual risk comparisons [48–50]. It also offers users the option to view a 2-minute visualization incorporating their avatar, previously developed by our team to illustrate how community immunity (herd immunity) may contribute to protection [44,45]. The visualization makes use of four theories or frameworks: the Health Belief Model [51], Gestalt visual principles [52], the Cognitive Theory of Multimedia Learning [53,54], and the Affect Heuristic [55,56]. We also added a values clarification feature, as per International Patient Decision Aid Standards, to help users align their vaccine decision with their values [57,58]. Finally, we used accordion menus (menus that show a brief title and allow people to click to open for full information) to enable users to decide for themselves how much or how little information they wish to read about the available vaccines [59]. We built the prototype in Vue.js (HTML, CSS, JavaScript) [60] with JSON-templated content. This architecture enabled rapid updates as new vaccine data and guidelines evolved.

Next, we conducted a third round of online user testing, again recruiting new participants (n=3) via tailored Facebook ads. Through individual semi-structured interviews on Zoom, we used think-aloud protocols focused on multi-device compatibility, navigation, and overall usability (technical details in Appendix 5).

Following user testing, the lead author (DE) presented the prototype at the 18th Biennial European Conference of the Society for Medical Decision-Making during the SMDM Core Course: Introduction to shared decision-making and patient decision aids. During the session, attendees engaged in a discussion in which attendees shared insights from their respective fields. As part of this discussion, the lead author (DE) had the opportunity to show the attendees the progress of our work and to get their feedback. This additional expert feedback (n=5) addressed some direction on medical content and interface navigation. In addition, an external user experience designer (n=1) delivered targeted feedback post-testing to complement the internal designer’s expertise.

While this cycle was underway, evolving public health guidelines suggested four distinct vaccination decisions: initial COVID-19 vaccination for those aged 12 years and above, booster doses for those aged 12 years and above, initial vaccination for children aged 6 months to 11 years, and booster doses for children aged 6 months to 11 years. We therefore restructured VaxDA-C19 accordingly.

#### Cycle 4

In Cycle 4, we finalized VaxDA-C19 such that it might be ready for testing in an online randomized controlled trial. We updated its content to reflect the most recent Health Canada-authorized COVID-19 vaccines and national recommendations from the National Advisory Committee on Immunization. We then invited all team members to comment on VaxDA-C19. Nine experts (n=9) provided detailed feedback. Evaluation criteria included scientific accuracy, linguistic clarity, completeness of content, and the comprehensive user journey across all four vaccination decisions.

Based on this feedback, we prioritized key modifications, including removing irrelevant vaccine selection options for specific age groups, clarifying instructional text to specify required selections more clearly, incorporating contact information and funder logos on the homepage, and resolving layout inconsistencies, notably mobile interface challenges.

Table 1 provides an overview of each cycle, along with the major modifications we implemented.

**Table 1.** Cycle Summaries.

| Cycle | Objective | Sample size (n) | Procedure used | Main modifications |
| --- | --- | --- | --- | --- |
| 1 | Needs assessment | n=9 users | Semi-structured interviews<br>Thematic analysis<br>Comparative testing | Clarified initial content |
| 2 | Intermediate refinement | n=22 users | Google Slides mock-ups<br>Comparative testing<br>Thematic analysis | Added avatar, accordion, icon arrays, visualizations, values clarification feature |
| 3 | Comprehensive usability testing | n=3 users + 6 experts | Think-aloud protocols<br>Functional prototype testing | Multi-device optimization<br>Refined navigation<br>Specified four distinct vaccination decisions |
| 4 | Scientific & bilingual review | n=9 experts | Expert review<br>User journey analysis | Updated content<br>Included French translation<br>Improved interface and instructional text |

##### Box 1.

###### Key points from the design and development process

**Recruitment strategies:** Recruitment through tailored social media advertisements (Facebook), complemented by internal dissemination via research team networks, effectively reached a diverse group of participants, including variations in educational level, age, language, and geographic location. Future studies could potentially benefit from additional strategies, such as direct collaboration with community-based organizations.

**Iterative usability testing:** Short, iterative usability cycles (four cycles, sample sizes from 3 to 22 participants each) allowed rapid identification and correction of usability issues. Participants’ repeated feedback emphasizes the need to simplify interactive elements (e.g., avatar creation, icon arrays, and value clarification sliders), which may have contributed to enhanced clarity and ease of use across testing cycles.

**Flexible content management:** Implementing JSON-based templates allowed efficient updates to rapidly-evolving vaccine guidelines, easing the task of ensuring ongoing scientific accuracy.

Early involvement of content experts contributed to the accuracy, clarity, and scientific rigor of the web-based patient decision aid.

**Multidisciplinary input:** Regular feedback from diverse interested parties (patients, healthcare professionals, user experience designers, and researchers) contributed a wide range of perspectives on the clinical relevance, usability, and accuracy of VaxDA-C19.

## Results

### Participant characteristics

Across the cycles, 45 participants responded to our user testing recruitment efforts, of which 34 completed one iterative user testing phase (cycle 1: n=9, cycle 2: n=22, cycle 3: n=3). Prospective participants who did not complete the testing did so because they did not wish to finish the study (n=2), were unavailable for any of the times scheduled for testing (n=4), responded after we had already reached saturation in findings (n=2) or for other reasons (n=3). Among the 34 participants, the mean age was 49 years (SD 16 years). The majority of our study participants were women, Canadian-born, and English speakers. Thirty-two percent of participants reported an ethnocultural identity other than white. Detailed demographics are shown in Table 2.

**Table 2:** User testing participant demographics across testing cycles (N=34)

| Category | Cycle 1 (n=9) | Cycle 2 (n=22) | Cycle 3 (n=3) |
| --- | --- | --- | --- |
| **Age (years), mean (SD)** | 47 (16) | 50 (16) | 52 (21) |
| **Birthplace: n (%)** |  |  |  |
| - Canada | 7 (78) | 21 (95) | 3 (100) |
| - Other | 2 (22) | 1 (5) | 0 (0) |
| **Preferred language(s): n (%) (multiple answers allowed)** |  |  |  |
| - French | 6 (67) | 5 (23) | 0 (0) |
| - English | 3 (33) | 18 (82) | 3 (100) |
| - Other | 0 (0) | 0 (0) | 0 (0) |
| **Ethnocultural group(s): n (%) (multiple answers allowed)** |  |  |  |
| - Indigenous or aboriginal person from outside of Canada (e.g., Native American, Maori, Quechua...) | 0 (0) | 0 (0) | 0 (0) |
| - Asian – East (e.g., Chinese, Japanese, Korean) | 0 (0) | 3 (14) | 0 (0) |
| - Asian – Central (e.g., Kazakhstani, Uzbekistani) | 0 (0) | 0 (0) | 0 (0) |
| - Asian – South (e.g., Indian, Pakistani, Sri Lankan) | 0 (0) | 1 (5) | 0 (0) |
| - Asian – South-East (e.g., Malaysian, Filipino, Vietnamese) | 0 (0) | 0 (0) | 1 (33.3) |
| - Black – African (e.g., Ghanaian, Kenyan, Somali) | 2 (22) | 1 (5) | 0 (0) |
| - Black – Caribbean (e.g., Barbadian, Jamaican) | 0 (0) | 2 (9) | 0 (0) |
| - Black – North American (e.g., Canadian, American) | 0 (0) | 0 (0) | 0 (0) |
| - First Nations | 0 (0) | 0 (0) | 0 (0) |
| - Inuit | 0 (0) | 0 (0) | 0 (0) |
| - Latin American (e.g., Chilean, Mexican, Salvadorian) | 0 (0) | 0 (0) | 0 (0) |
| - Metis (Metis Nation in Canada) | 0 (0) | 1 (5) | 0 (0) |
| - Middle Eastern (e.g., Egyptian, Iranian, Lebanese) | 0 (0) | 0 (0) | 0 (0) |
| - North African (e.g. Moroccan, Tunisian) | 0 (0) | 0 (0) | 0 (0) |
| - White/European (e.g., English, Italian, Portuguese, Russian) | 1 (11) | 6 (27) | 1 (33.3) |
| - White/North American (e.g., Canadian, American) | 6 (67) | 10 (45) | 1 (33.3) |
| - Other | 0 (0) | 0 (0) | 0 (0) |
| - Prefers not to answer | 0 (0) | 0 (0) | 0 (0) |
| **Disability status: n (%)** |  |  |  |
| - No disability | 7 (78) | 15 (68) | 3 (100) |
| - At least one disability | 2 (22) | 6 (27) | 0 (0) |
| - Prefers not to answer | 0 (0) | 1 (5) | 0 (0) |
| **Gender identity: n (%)** |  |  |  |
| - Woman | 7 (78) | 12 (55) | 3 (100) |
| - Man | 2 (22) | 10 (45) | 0 (0) |
| - Indigenous or other cultural gender minority identity (e.g., two-spirit) | 0 (0) | 0 (0) | 0 (0) |
| - Something else (e.g. non-binary, gender-fluid) | 0 (0) | 0 (0) | 0 (0) |
| - Prefers not to answer | 0 (0) | 0 (0) | 0 (0) |
| **Highest education level completed: n (%)** |  |  |  |
| - Some elementary school (completed or not) | 0 (0) | 0 (0) | 0 (0) |
| - High school diploma | 1 (11) | 2 (9) | 0 (0) |
| - Apprenticeship or trade certificate or diploma | 0 (0) | 0 (0) | 0 (0) |
| - College or polytechnical school certificate or diploma | 1 (11) | 7 (32) | 0 (0) |
| - University degree, bachelor level or below | 3 (33) | 10 (45) | 2 (67) |
| - University graduate degree (Master's or Doctorate level) | 4 (44) | 2 (9) | 1 (33) |
| - Prefers not to answer | 0 (0) | 1 (5) | 0 (0) |
| - Do not know | 0 (0) | 0 (0) | 0 (0) |
| **Sexual orientation: n (%)** |  |  |  |
| - Straight or heterosexual | 8 (89) | 17 (77) | 3 (100) |
| - Lesbian, gay or homosexual | 1 (11) | 1 (5) | 0 (0) |
| - Other (options included Bisexual, Something else, and Prefers not to answer) | 0 (0) | 4 (18) | 0 (0) |
| **Health status: n (%)** |  |  |  |
| - Poor | 1 (11) | 2 (9) | 0 (0) |
| - Fair | 0 (0) | 3 (14) | 0 (0) |
| - Good | 2 (22) | 8 (36) | 0 (0) |
| - Very Good | 5 (56) | 7 (32) | 2 (67) |
| - Excellent | 1 (11) | 2 (9) | 1 (33) |
| **Public facing occupation: n (%)** |  |  |  |
| - Yes | 2 (22) | 5 (23) | 0 (0) |
| - No | 7 (78) | 16 (73) | 3 (100) |
| - Prefers not to answer | 0 (0) | 1 (5) | 0 (0) |
| **Provinces and Territories: n (%)** |  |  |  |
| - Alberta | 1 (11) | 0 (0) | 0 (0) |
| - British Columbia | 0 (0) | 1 (5) | 0 (0) |
| - Manitoba | 0 (0) | 4 (18) | 0 (0) |
| - New Brunswick | 0 (0) | 1 (5) | 0 (0) |
| - Newfoundland and Labrador | 0 (0) | 0 (0) | 0 (0) |
| - Nova Scotia | 0 (0) | 0 (0) | 0 (0) |
| - Ontario | 2 (22) | 11 (50) | 3 (100) |
| - Prince Edward Island | 0 (0) | 0 (0) | 0 (0) |
| - Quebec | 6 (67) | 5 (23) | 0 (0) |
| - Saskatchewan | 0 (0) | 0 (0) | 0 (0) |
| - Northwest Territories | 0 (0) | 0 (0) | 0 (0) |
| - Nunavut | 0 (0) | 0 (0) | 0 (0) |
| - Yukon | 0 (0) | 0 (0) | 0 (0) |
| **Residing in a rural community: n (%)** |  |  |  |
| - No | data not available | 17 (77) | 2 (67) |
| - Yes | data not available | 2 (9) | 1 (33) |
| - Don't know | data not available | 3 (14) | 0 (0) |

**Table 3.** Summary of thematic feedback from Cycle 1 user testing (n=9) and changes made following citizen panel recommendations.

| Theme | Key Feedback/Issue | Participants<br>n (%) | Resulting modifications |
| --- | --- | --- | --- |
| **Avatar arrays** | Interpreted as a “group of people”; needed a clearer explanation that each icon represents one individual in a population | 9 (100) | Added explanatory text indicating probabilistic meaning |
| **Icon array design preferences** | Circular array for better spacing | 5 (56) | Refined visual layout, used more spacing |
|  | Shirt-colored array for character detail | 3 (33) | None (optional alternate design retained for future testing) |
| **Vaccine question framing** | Neutral feelings about the question | 5 (56) | Adjusted question to more neutral wording |
|  | Would consult a healthcare provider | 2 (22) | None; consistent with decision aid concept |
|  | Increased reassurance | 1 (11) | None |
|  | Felt pressured, uncomfortable | 1 (11) | Wording revised for less prescriptive tone |
| **Vaccine explanation text** | Effective explanation of rapid vaccine development | 9 (100) | Retained main points; reorganized structure |
|  | Perceived as too technical/lengthy | 5 (56) | Simplified text and terminology |
|  | Reassuring | 3 (33) | Maintained neutral tone |
|  | Confusion with the first sentence | 3 (33) | Clarified opening sentence |
|  | Insufficient details on side effects | 1 (11) | Emphasized side-effect transparency in revised draft |

### Cycle 1

#### Main findings

##### Icon-array interpretation

All participants (n=9) correctly identified the icon arrays as representing a “group of people” and understood that the colored icons indicated numerical or probabilistic information. However, some participants struggled initially with the precise probabilistic meaning of each icon. Regarding layout preferences, five participants preferred the arrays using circular backgrounds, finding them “easier to count” due to clearer spacing and structure. Three participants favored the shirt-colored arrays because they appreciated seeing “more character detail.” One participant found both layouts equally clear but mentioned that the shirt-colored icons seemed to visually represent more data because of their elongated shape.

##### Provider-sent message framing

When participants reacted to the hypothetical vaccine recommendation message from their primary care provider (Appendix 1), five participants (5/9) expressed neutral feelings toward the message. Two participants (2/9) indicated that receiving such a message would encourage them to consult their provider with further questions. One participant (1/9) found the message reassuring, whereas another (1/9), who had expressed prior vaccine concerns, felt “pressured and uncomfortable.”

##### Vaccine-confidence social media text

All participants (9/9) appreciated the explanation provided about the rapid vaccine development process (Appendix 2). Nevertheless, several participants raised concerns regarding the text’s accessibility and readability. Specifically, five participants (5/9) thought the text was appropriate mainly for highly educated people or those who already trust the authorities. Three participants (3/9) indicated that the text was too long. Three participants (3/9) found this message reassuring. Another three participants (3/9, 1 English-speaking, 2 French-speaking) reported confusion with the opening sentence, unsure how it related to the subsequent content. Additionally, one participant (1/9) expressed concerns about insufficient transparency regarding potential vaccine side effects.

##### Citizen panel recommendations

The citizen panel (n=6) recommended incorporating relatable real-world data into the patient decision aid, automating vaccine selections based on provincial guidelines, and helped to refine visual branding with a logo.

#### Changes for the next cycle

Based on these findings, we implemented several targeted modifications to improve our patient decision aid. We explicitly clarified that each icon represented one individual in a given population to address confusion around probabilistic visualization. Considering mixed feedback on icon-array layouts, we retained both designs for further testing.

We revised the healthcare provider message to a more neutral tone, addressing concerns about pressure or discomfort. Finally, to improve the vaccine-confidence content, we reorganized the structure to enhance clarity. We simplified technical language, clarified the opening sentence, and incorporated more transparent information about potential side effects, in line with participants’ expectations regarding tone, accessibility, and completeness.

### Cycle 2

#### Main findings

During this second round of user testing, participants (n=22) reviewed two content variants designed to explain the rapid development of COVID-19 vaccines. The objective was to assess the clarity, effectiveness, and comprehensiveness of each version. Version A contained only illustrations, while Version B included photographs and researchers’ names. We randomly assigned 11 participants to Version A and 11 to Version B. Participants answered questions on message interpretation and identified important unanswered questions about COVID-19 vaccination missing from the explanations.

##### Reactions to the VaxDA-C19 team presentation

Participants expressed polarized opinions about Version B. Two participants appreciated the inclusion of researchers’ names and photos, interpreting it as a sign of credibility and transparency. One participant remarked that it was “good to see a Manitoba doctor” involved, while another said that the real faces “gave some credibility”. Others expressed suspicion and discomfort, perceiving the design as overly promotional or politically biased. Participants describe the content using terms like “political PR,” “spin”, or sarcastic phrases such as “a miracle that will solve everything”. Several participants misinterpreted the visuals, believing these scientists developed the vaccines. Some participants perceived the section as an attempt to distance pharmaceutical companies from the vaccine’s origins, and described this as reassuring or misleading.

##### Interpretation of vaccine explanation text

In response to the question, “Can you tell me what this text was about?”, most participants described an explanation of how vaccines were developed (Version A: 7/11, Version B: 9/11). Five participants in each group thought the section highlighted the scientists behind the vaccine development. A minority said the message aimed to counter vaccine skepticism or clarify funding sources. Table 4 summarizes how participants interpreted the vaccine explanation text.

**Table 4.** Cycle 2 user testing participants’ thematic understanding of vaccine explanation text (n=22)

| Themes | Feedback | Version A (n=11)<br>n (%) | Version B (n=11)<br>n (%) | Action taken |
| --- | --- | --- | --- | --- |
| Understanding | Clear explanation of the vaccine development process | 7 (64) | 9 (82) | Retained detailed version |
| Interpretation | These are the scientists who worked on the vaccines | 5 (46) | 5 (46) | Clarified scientist roles |
| Interpretation | Text aims to ease anxieties or respond to anti-vaccine concerns | 3 (27) | 4 (36) | Retained supportive messaging |
| Interpretation | Vaccine research is **not** funded by pharmaceutical companies | 2 (18) | 2 (18) | Clarified funding explicitly |
| Interpretation | None of the above | 1 (9) | 1 (9) |  |

##### Reactions to the vaccine explanation text

During this round of testing, participants (n=22) provided varied emotional reactions and interpretations regarding the explanatory text on rapid COVID-19 vaccine development. Two participants perceived a clear division between the scientists or authorities and the public, illustrated by expressions of suspicion or detachment such as “The people who created it are vouching for it,” and referring to authorities as “Them.”

Some participants identified important information gaps, particularly the lack of details on vaccine effectiveness, side effects and characteristics of clinical trial participants. For instance, a participant remarked that the vaccine was presented as “a miracle that will solve everything”, while another expressed nuanced skepticism: “I know exactly how it’s done, and I take some and leave some […] I’m a bit perplexed”. Two participants specifically indicated confusion or misunderstanding about the sentence stating, “Other vaccines are still being tested in places with fewer COVID-19 cases.” One described needing more time to understand the phrase clearly, while the other explicitly found it ambiguous.

One participant strongly criticized the text as too political and not scientific, disputing claims about vaccine availability and the accuracy of funding statements. Another felt the text insufficiently emphasized the societal consequences of refusing vaccination, remarking strongly that, “It’s basically a crime if people are not taking this seriously.”

Finally, one participant noted perceived contradictions within the explanation, particularly regarding how previous coronavirus research aligns with the rapid vaccine development and funding during the pandemic. This participant also highlighted inconsistencies in public health messages from different provincial health authorities. Table 5 below summarizes the main emotional reactions and comprehension issues identified.

**Table 5.** Summary of emotional reactions and information gaps.

| Theme | Key issues | Example quotes | Frequency |
| --- | --- | --- | --- |
| **Detachment or suspicion** | Authorities perceived as separate or untrustworthy entities | “The people that made it are vouching for it.”<br>““Them” trying to explain[...], “in ‘their’ opinion, [a] vaccine is a source of cure.” | 2/22 |
| **Information gaps** | Lack of clarity on vaccine effectiveness, side effects, and allergens | “Now it's presented as a miracle that will solve everything.”<br>“I'm a bit perplexed.” | ≥2/22 |
| **Confusion or ambiguity** | Misinterpretation of the statement about vaccine testing locations | “Would need more time to understand this one.”<br>“I find the last sentence a little ambiguous.” | 2/22 |
| **Political or promotional tone** | Perceived political bias and promotional language | “Canada has a ton of vaccines!” | 1/22 |
| **Societal consequences emphasis** | Insufficient focus on consequences of non-vaccination | “It's basically a crime if people are not taking this seriously.” | 1/22 |
| **Perceived contradictions** | Confusion about consistency and accuracy of provided information | “Scientists have been working on the coronavirus since 2003 [...] It's not new. [...] Even health ministers from different provinces say different things.” | 1/22 |

##### Unanswered questions

Table 6 summarizes the types of questions participants felt remained unanswered after reading the vaccine explanation text. These focus on vaccine mechanisms, COVID-19-specific information, distribution and eligibility, safety, and research.

**Table 6.** Themes of unanswered questions (n=22)

| Theme | Feedback | n (%) |
| --- | --- | --- |
| **General mechanisms and effects** | How does the vaccine work? What does it do to the virus? Why two doses? What does it do to your body? Are there side effects? | 9 (41) |
| **COVID-19 vaccine-specific information** | What is this technology? Why are there different vaccines? How were they made, what's in them? What are variants and do the vaccines cover them? What protection does it give? How long before it kicks in? If I get COVID-19, does it mean I don't need a vaccine? Is it like the flu vaccine that you should get each year? | 12 (55) |
| **Distribution and eligibility** | How are they distributed, when can we get it? Who should and shouldn't get the vaccine? What happens if you don't get the vaccine? | 7 (32) |
| **Safety and research** | Is the vaccine safe? Were they tested enough? Is there enough data? Were people with different conditions part of the trials? | 6 (27) |
| **No remaining questions** | - | 2 (9) |
| **Question not asked** | - | 1 (5) |

#### Changes for the next cycle

Participants feedback in Cycle 2, we implemented several changes to improve clarity, transparency, and user trust in the next iteration of VaxDA-C19 decision aid. We moved the photos and names of scientists to a dedicated “About Us” section to reduce perceptions of promotional content and clarified their roles to reduce confusion. While keeping the explanatory text detailed, we simplified complex terms to increase accessibility. In response to recurring concerns and unanswered questions about vaccine technology, safety, variants, and side effects, we also expanded the frequently asked questions (FAQs) section.

### Cycle 3

#### Main findings

During Cycle 3, we conducted a third round of user testing (n=3) to assess navigation, usability, and comprehension across devices. Table 7 presents a thematic summary of participant feedback.

**Table 7.**
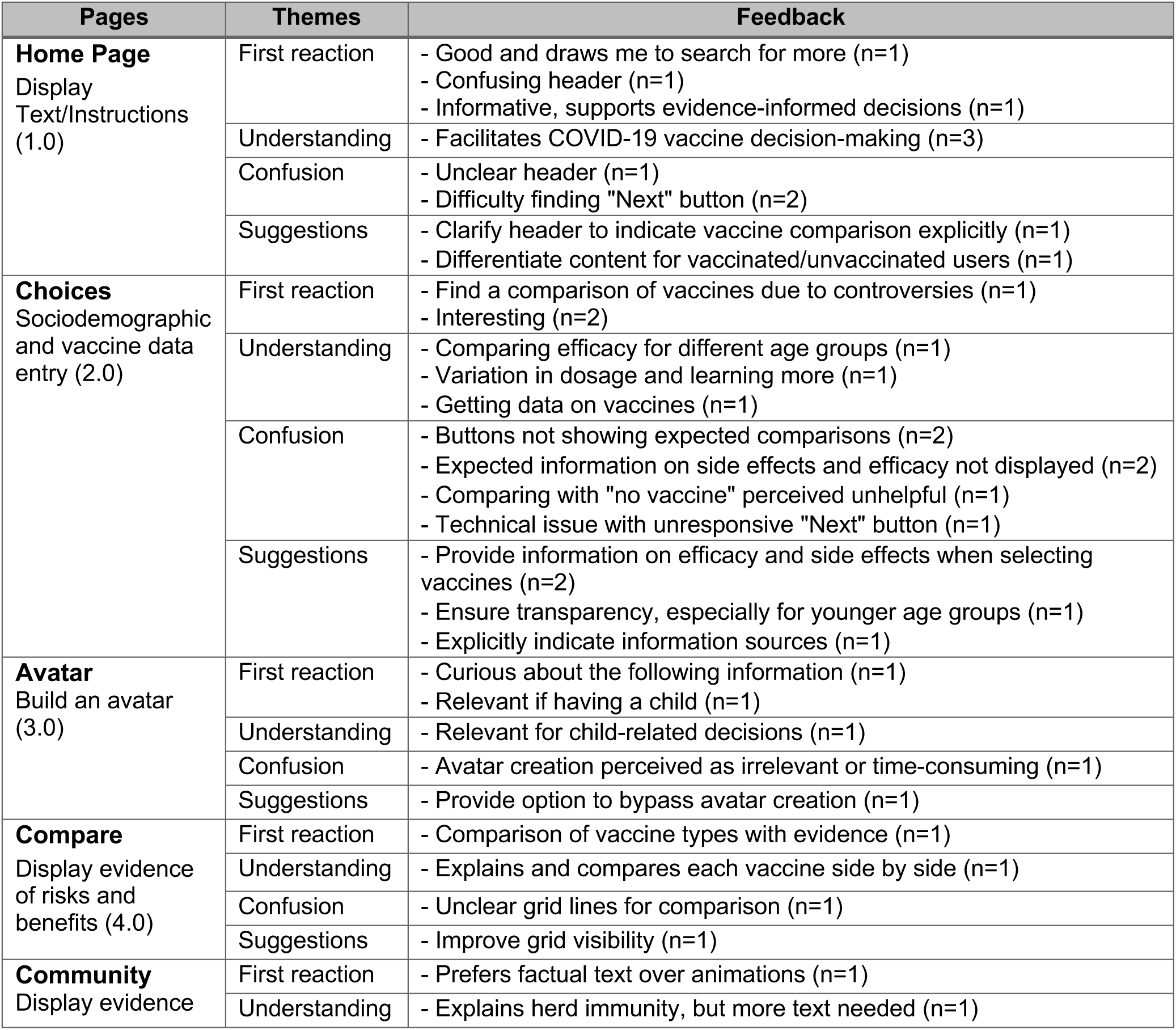
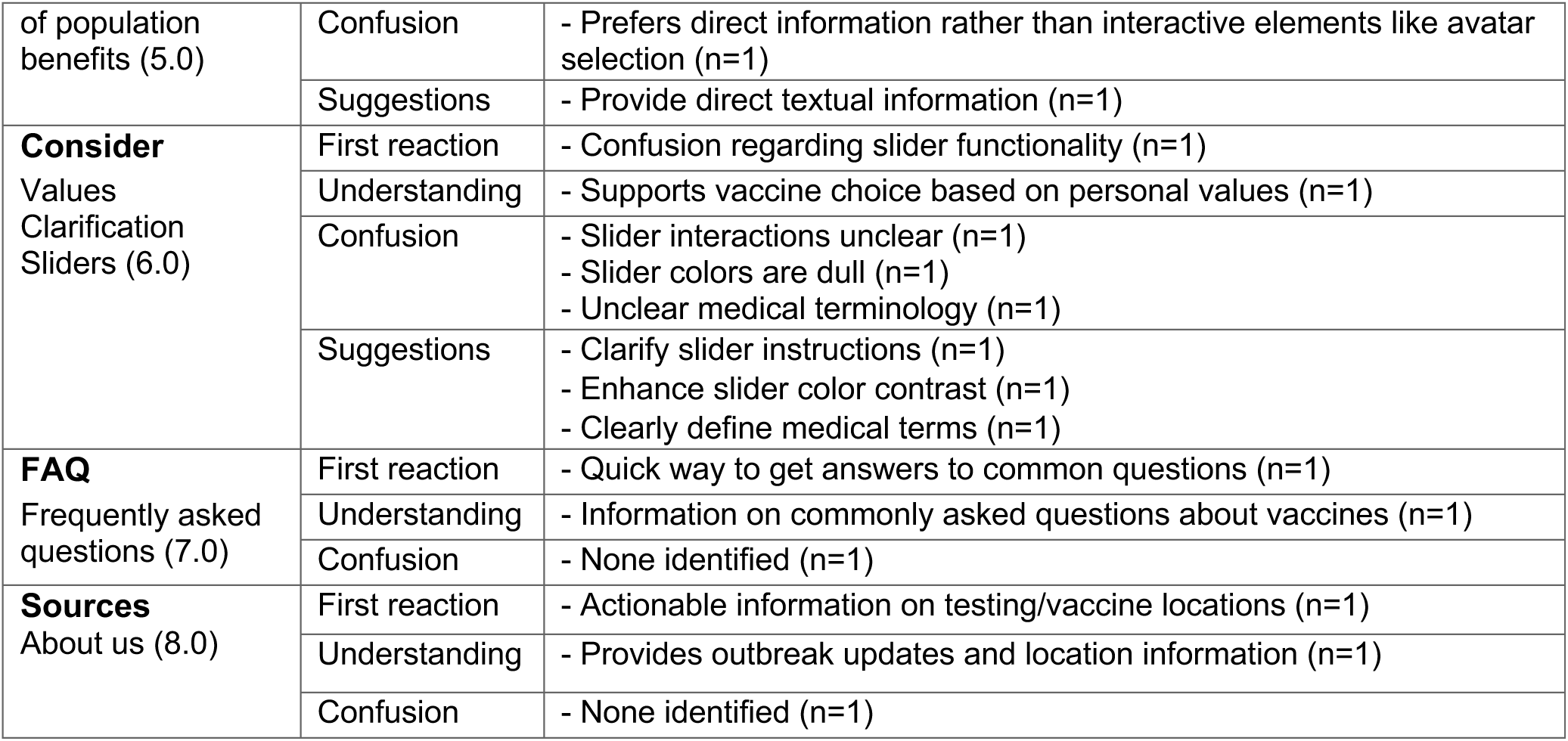
Summary of thematic feedback from Cycle 3 user testing (n=3)

##### Navigation and usability improvements

Participants (n=3 users, 6 experts) identified significant navigation issues, notably difficulties finding the “Next” button, which impaired smooth transitions between sections. They also recommended clearer labelling and explicit instructions to clarify interactions with pages involving comparative vaccine selection and slider functionalities.

##### Information transparency and accessibility

Participants emphasized the need for simplified medical terminology and a more transparent presentation of vaccine efficacy and side effects data. They noted that clear, immediate comparative data would be crucial, particularly important for parents evaluating COVID-19 vaccination for children.

##### Interactive features optimization

Interactive elements, such as avatar creation and values clarification sliders, received mixed responses. Participants initially perceived avatar creation positively, but one participant found creating multiple avatars to form a community as unnecessary or excessively time-consuming. One participant had a preference for factual information rather than interactive elements or animations. Sliders were occasionally confusing due to unclear functionality interactions, dull colors and poorly defined medical terms.

##### Insights from external experts

Five healthcare professionals and medical decision-making researchers (n=5) provided feedback at a medical-decision making conference. This feedback included recommendations for improved mobile navigation, structured vaccine selection menus, refined content spacing, and modal placements. A subsequent external reviewer (n=1) with expertise in user experience recommended adding a step-by-step user guide and informative pop-ups to support user interactions.

#### Changes for the next cycle

Following Cycle 3, we implemented targeted modifications to improve navigation, content clarity, and overall user experience in VaxDA-C19. We enhanced button visibility and repositioned key navigation elements, adding clear user instructions to support a smoother transition. To increase content accessibility and transparency, we simplified medical terminology and added comparative data on vaccine efficacy data and side effects directly within relevant sections. To streamline interactivity, we introduced an avatar bypass option and clarified the functionality of values clarifications sliders through revised instructions and enhanced visual design. We also applied external expert recommendations to improve mobile navigation, refine spacing, structure vaccine menus more clearly, and integrate contextual guidance through informative pop-ups.

### Cycle 4

#### Main findings

Cycle 4 consisted of a comprehensive expert review (n=9), focusing on usability, clarity, bilingual implementation, and overall application refinements as shown in Table 8.

**Table 8.** Cycle 4: Expert review feedback and implemented modifications.

| Section | Expert feedback | Priority | Implemented modifications |
| --- | --- | --- | --- |
| **Avatar creation** | Improve logical flow | High | Repositioned avatar creation |
| **Decision clarity** | Clarify vaccine selection instructions | High | Added explicit instructional texts |
| **Decision clarity** | Include the "I don't know" option clearly | High | Option clearly added |
| **Mobile interface** | Resolve text overlap and navigation issues | High | Improved responsive design and optimized mobile usability |
| **Language** | Complete bilingual implementation | High | Fully implemented bilingual interface (English/French) |
| **Content clarity** | Clarify wording of decision context (e.g., "I am deciding about...") | High | Decision wording clarified consistently across all steps |
| **Community immunity animation** | Adjust slow-running animations, clarify instructions, add a skip function | High | Animation optimized, explicit instructions provided, skip function added |
| **Values clarification sliders** | Address confusion around slider instructions and colors | High | Instructions repositioned clearly above sliders, colors adjusted for clarity |
| **Avatar usability** | Clarify the avatar skipping process | High | Clear option added to bypass avatar creation easily |
| **Navigation** | Allow free navigation across sections via the bottom path bar | Medium | Navigation bar updated for easier movement between sections |
| **Home page** | Display funder/institutional logos and contact information | Medium | Logos and contact details added clearly on the homepage and FAQ |
| **Visual layout** | Improve spacing of provincial selection buttons | Low | Implemented in the final version |
| **Icon arrays** | Resolve color confusion with face icons | Low | Not implemented (consideration for future visual redesign) |
| **Community immunity section** | Add customizable animations speed/pause options | Low | Not implemented (evaluated for technical complexity in future versions) |
| Information density | Reduce textual density | Low | Not implemented (noted for future text optimization) |
| Vaccine comparison | Simplify side-by-side visual vaccine comparison visuals difficult | Low | Not implemented (requires future technical exploration) |

##### High-priority usability improvements

Experts (n=9) identified several issues affecting clarity and user experience. These included irrelevant vaccine comparisons for children under 12, unclear user instructions, confusing avatar placement, navigation difficulties on the mobile interface, and incomplete French implementation. Experts also suggested clarifying the decision-making context consistently across the web-based application (e.g., clearly indicating options such as “I don’t know”) and recommended optimizations for animations and interactive sliders to reduce confusion and enhance usability.

##### Moderate-to-low priority feedback

Additional recommendations included improving free navigation across sections, clearly displaying institutional logos and contact information, enhancing visual spacing for provincial selection buttons, and refining icon-array color schemes. Experts also proposed custom speed and pause features for animations, reducing overall text density, and simplifying vaccine comparison visuals. Due to technical and feasibility constraints, we recorded moderate-to-low priority feedback for future updates.

#### Implementation of the final version of VaxDA-C19

VaxDA-C19 evolved from an initial Google Slides mock-up into a fully functional, bilingual web-based application. Key technical improvements include transitioning to a responsive HTML5 and JavaScript architecture, with real-time content synchronization. Major features include automated visualizations for herd immunity, refining comparative interactive features, and enhanced icon arrays. These refinements ensure that VaxDA-C19 remains human-centred, responsive to evolving vaccination guidelines, while improving usability and user trust.

##### Box 2.

###### Key points from usability testing results

**Navigation and interface usability:** Participants consistently highlighted the need for clear, intuitive, and easily navigable interfaces. Regular usability checks using small iterative samples allowed the research team to quickly identify critical navigation barriers. Participants recommended explicit labelling of navigation buttons (such as the “Next” button), logical repositioning of interactive elements, and improved visibility of control. These adjustments may improve user satisfaction and reduce cognitive load.

**Information transparency and accessibility:** Participants frequently emphasized the importance of clear, concise, and transparent information on vaccine efficacy and side effects. The research team anticipated that simplifying medical language and providing immediate comparative information could enhance user engagement, comprehension, and trust. Participants identified addressing transparency around sensitive health decisions, such as pediatric vaccination, as important to increase the usability and credibility of the patient decision aid.

**Interactive features optimization:** Participants suggested explicitly defining medical terms, clarifying instructions for using interactive elements (avatars, values clarification sliders), optimizing slider functionalities, and offering optional bypass features for less non-essential interactive steps. The research team incorporated these recommendations to reduce perceived cognitive burden and potentially enhance user experience, though further validation remains necessary.

**Responsive content adjustments:** User feedback identified specific gaps in information (e.g., vaccine efficacy, technology, clinical trial, and distribution logistics) and highlighted areas susceptible to misunderstanding or skepticism (e.g., funding sources, researchers’ roles). Participants recommended expanding and refining frequently asked questions (FAQs), clearly explaining scientific roles, and transparently communicating funding and clinical processes. The research team implemented these content adjustments to address user skepticism, improve comprehensiveness, and enhance perceived relevance.

**Multidisciplinary and expert reviews:** Feedback from multidisciplinary perspectives, including healthcare professionals, user experience specialists, and citizen panels, helped refine the web-based patient decision aid’s usability and scientific content. The research team prioritized and implemented expert recommendations (e.g., improved mobile navigation, structured vaccine selection, clear scientific explanations). These refinements aimed to enhance clarity, accessibility, and user acceptance, although further formal validation of their impact is required.

## Discussion

### Principal Findings

This study aimed to iteratively develop and refine a bilingual (English and French) web-based patient decision aid (VaxDA-C19) to help people in Canada make evidence-informed, values-congruent decisions about COVID-19 vaccination. VaxDA-C19 includes interactive components such as customizable avatars, personalized icon arrays constructed out of those avatars, an optional, animated, theory-based visualization of community immunity, and a values clarification method. We included these features based on their capacity to facilitate effective communication, optimize cognitive load, enhance empathy, and provide clear, evidence-based information on vaccine options, benefits, and risks [32,44,45,48,57–59]. We present three principal findings from this work.

First, a recurrent theme in participant feedback was the need for more detailed and transparent information about vaccine efficacy, variant coverage, immunity duration, and the inclusivity of clinical trials. To address these concerns, enhance transparency and hopefully reinforce trust in the information presented, rather than offering a brief list of references in a separate section at the end of the document or website, as is commonly done in patient decision aids, we integrated explicit links to peer-reviewed studies and health authority documents immediately following each statement. We used the word “citation” for each reference and linked directly to open-access resources whenever possible. We also designed visual strategies, such as including images of scientists, to increase credibility [61–63]; however, responses to this approach varied. Some participants found this approach reassuring, while others perceived it as manipulative or politically motivated. To ultimately mitigate these concerns, we removed images and prioritized emotional neutrality and transparent, nonjudgmental communication [1,4,15,64–66]. This approach aligns with the Trust Determination Theory, which emphasizes how credibility, inclusivity, and clarity foster public confidence and reduce skepticism [67]. Similarly, the Extended Parallel Process Model (EPPM) highlights how fear, distrust, and perceived governmental or institutional biases influence message reception, potentially diminishing the effectiveness of health communication [68].

Second, rather than seeking an elusive one-size-fits-all information presentation, presenting a simple overview of essential points and allowing people to easily access more detail when they want it may better serve a wider range of information needs. People with lower health literacy, who often express greater vaccine hesitancy, find more transparent explanations of risks and benefits particularly helpful [64,69–71]. Some participants in our study similarly emphasized the importance of clear, simplified explanations, particularly for individuals with varying levels of health literacy, and also for people who lack the time or interest to delve deeply into information about vaccines. However, as noted above in our first point, other participants wanted far more detail. These tensions between some people’s desires for comprehensive information and others’ desires for more succinct information highlighted the complexity inherent in balancing simplicity and visual appeal with informational accuracy [72,73]. To address these competing priorities, we revised medical terminology, restructured content for better readability, and ultimately presented a summary of key evidence to all users, followed by accordion menus, which allow users to expand sections to read more about a given topic. By giving people control over the amount of detail they receive, we hope VaxDA-C19 will serve the needs of people who want a summary of essential points as well as people whose questions are not answered by simplified public health messages. Future research should continue to examine optimizing content presentation to provide sufficient detail without overwhelming users.[74]

Third and finally, specific challenges associated with developing a patient decision aid for COVID-19 vaccination included a novel pathogen that launched a global pandemic, novel vaccine technology, continuously evolving evidence about vaccine effectiveness and adverse events following immunization, changing vaccine products and authorizations, and evolving recommendations for use. The fast-evolving COVID-19 vaccination landscape required proactive content management. We used flexible structures like JSON templates to enable timely updates aligned with new evidence and public health recommendations. This technical adaptability, combined with sufficient time commitments on the part of team members, should help us ensure that the VaxDA-C19 will remain scientifically accurate, while also facilitating version control, scalability, and potential future reuse for patient decision aids about other vaccine-preventable diseases.

### Comparisons with existing literature

The COVID-19 pandemic led to the rapid development of digital patient decision aids aimed at addressing vaccine hesitancy, which intensified due to misinformation and information overload (infodemic) [75]. Several countries, including France [76], the Netherlands [77], and Australia [78], introduced digital tools to support informed decision-making. Compared to static informational approaches, these interactive decision aids improved user knowledge, reduced decisional conflict, and increased satisfaction with the decision-making process [75,77]. Despite these benefits, major challenges persist, including potential biases in content presentation, variations in complexity, and inconsistent accessibility, all of which affect usability and effectiveness [75,77].

International evaluations identified key factors that contribute to the success of digital patient decision aids. Co-design approaches that involve health professionals and target populations enhance relevance and usability. Regular updates ensure content accuracy and alignment with evolving public health guidelines. User-friendly interfaces and personalization based on individual risk assessments further support engagement and informed decision-making [75–78]. Certain digital patient decision aids have also integrated practical features, such as direct scheduling links for vaccination appointments, to facilitate action [75,77]. Similarly, in our study, multiple cycles of qualitative usability testing within an iterative, human-centered approach helped identify and address navigation, comprehension, and usability issues early in development. Small-scale usability tests effectively detected key issues, allowing for rapid refinements. Cognitive ergonomics literature supports this strategy, indicating that explanatory texts provide more explicit contextual cues and facilitate information processing, particularly for individuals with lower health literacy [79–81].

Limited linguistic and cultural adaptability restricts the reach of many existing digital patient decision aids, particularly among culturally and linguistically diverse populations [75,77]. VaxDA-C19 aims to address this limitation initially through its bilingual content in English and French, the official languages of Canada. Following evaluation in an online randomized controlled trial, we also plan to translate it into other major languages spoken in Canada.

Although digital patient decision aids may influence vaccine intentions, evidence directly linking these tools to increased vaccine uptake remains incomplete. Ongoing randomized controlled trials, such as SMART-DA in Korea [82], aim to assess their impact on key outcomes, including vaccine intentions, knowledge retention, decisional conflict, and emotional responses. Findings suggest that patient decision aids improve preventive health behaviors by enhancing decision quality rather than by applying persuasive messaging [83,84].

Our study aligned with previous research suggesting that effective communication in patient decision aids like VaxDA-C19 requires careful consideration of emotions, trust, and perceived biases [1,4,10,85]. Emotional responses play an important role in how individuals interpret vaccine-related messages [1,15]. In our study, some participants expressed skepticism, anxiety, or distrust, particularly if they perceived wording as being politically charged, overly promotional, ambiguous, or institutionally biased. This finding aligns with other studies [86] and these reactions overall align with Sandman’s Risk = Hazard + Outrage model, which posits that public outrage or mistrust can overshadow objective risk assessments, leading to defensive responses such as rejection or disengagement [4,18,86,87]. Pre-existing attitudes towards vaccines may also influence people’s reactions to interventions such as VaxDA-C19. Future research should incorporate such considerations into study design.

Our study also confirmed that intuitive navigation, visual clarity, and balanced content presentation played essential roles in usability. These findings align with existing literature, which demonstrates that interactive and personalized elements improve engagement and comprehension in digital health interventions [48,88,89].

## Strengths and limitations

Our study has two main limitations. First, while recruiting user testing participants through tailored Facebook advertisements allowed us to reach people living in different parts of Canada with diverse backgrounds and who are online and therefore potential users of a web-based patient decision aid, no recruitment platform can perfectly represent the broader population. We note that our user testing participants were, as a group, slightly older than the broader group of Facebook users in Canada. Specifically, among Facebook users in Canada, the largest age group (around 25%) is between 25 and 34 years old, with the next largest group (19%) being 35 to 44 years old [90]. We may have recruited more people older than these groups due to increasing interest in COVID-19 vaccines with age. The age group we recruited was approximately representative of the desired population for a patient decision aid about COVID-19 vaccines. Seniors in Canada were already receiving strong recommendations to obtain the vaccine, whereas people in the age range of our user testing participants were likely to be both candidates themselves and may also be parents of children who were candidates for COVID-19 vaccination at the time of the study. Our study participants also included more people with higher levels of education than the general population in Canada. People with more education may seek more information about health compared to people with less education. Second, the challenge of balancing comprehensive content with cognitive load remains potentially unresolved, particularly in adapting information for users with varying levels of health literacy. While brief, simple messaging can be most effective in some contexts and for some people, the resulting lack of detail and missing explanations can be a problem for others. Variability in patient decision aid formats, development methodologies, and user preferences suggests that no single approach fits all contexts [91,92]. We therefore chose to prioritize user control over information presentation, trusting that many people will know what they need. Nevertheless, additional research should investigate these aspects further to refine content presentation and improve the effectiveness of digital patient decision aids.

Our study also has two main strengths. First, we used a rigorous iterative design based on human-centered methodologies, a multidisciplinary approach, bilingual validation, and structured engagement of a range of interested parties, including user testing, a citizen panel, user experience interviews, and expert reviews. The collaboration between a diversity of researchers, user experience specialists, members of the public, and healthcare professionals helped refine the patient decision aid, contributing to its scientific reliability and usability. User testing participants from different backgrounds offered different responses and reactions, providing a range of feedback to which we were able to respond. Complementing the contributions of these members of the public who participated in user testing and saw VaxDA-C19 at a single time point, members of the citizen panel saw VaxDA-C19’s evolution over time and were able to comment at a higher level on how well or poorly the design was responding to its original goals as well as the evolving pandemic context. A wide range of experts brought insights from related projects, literature, guidelines, and also lent their expertise to verify VaxDA-C19’s content and design. Systematically integrating all these people’s feedback on an ongoing basis allowed for continuous adjustments. Second, we believe that VaxDA-C19 fills an unmet need by bridging the gap between highly technical vaccine information and highly simplified public health messaging. As COVID-19 vaccines continue to be offered to people in Canada, we hope to help support high-quality decision-making, thus contributing to the health of people in Canada, and to public health more broadly. The platform itself may also be adapted to other vaccines.

We plan to assess the effects of VaxDA-C19 in an online randomized controlled trial. Specifically, participants who are eligible to receive COVID-19 vaccines will be randomly assigned to one of three arms: a control group receiving no additional information, a usual information group directed to the Public Health Agency of Canada’s COVID-19 vaccination web pages, or an intervention group receiving access to VaxDA-C19. We will assess effects on the primary outcome of vaccination intentions and secondary outcomes: knowledge, emotions, decisional conflict, vaccine uptake, and decisional regret. We will also examine effects on trust in the information provided, comparing only between usual information and VaxDA-C19. Finally, we will conduct a follow-up study 3–6 months later to ascertain whether or not participants received COVID-19 vaccines.

## Conclusions

VaxDA-C19 is an online patient decision aid designed and developed to help people in Canada make evidence-informed, values-congruent decisions about COVID-19 vaccines on an ongoing basis. An iterative, human-centered design approach and a multidisciplinary team bringing diverse perspectives led to a flexible application that we hope will prove useful for years to come.

## Data Availability

All data generated or analyzed during this study are included in this published article and are available on demand.

## APPENDICES

### Appendix 1. User testing interview guide - Cycle 1

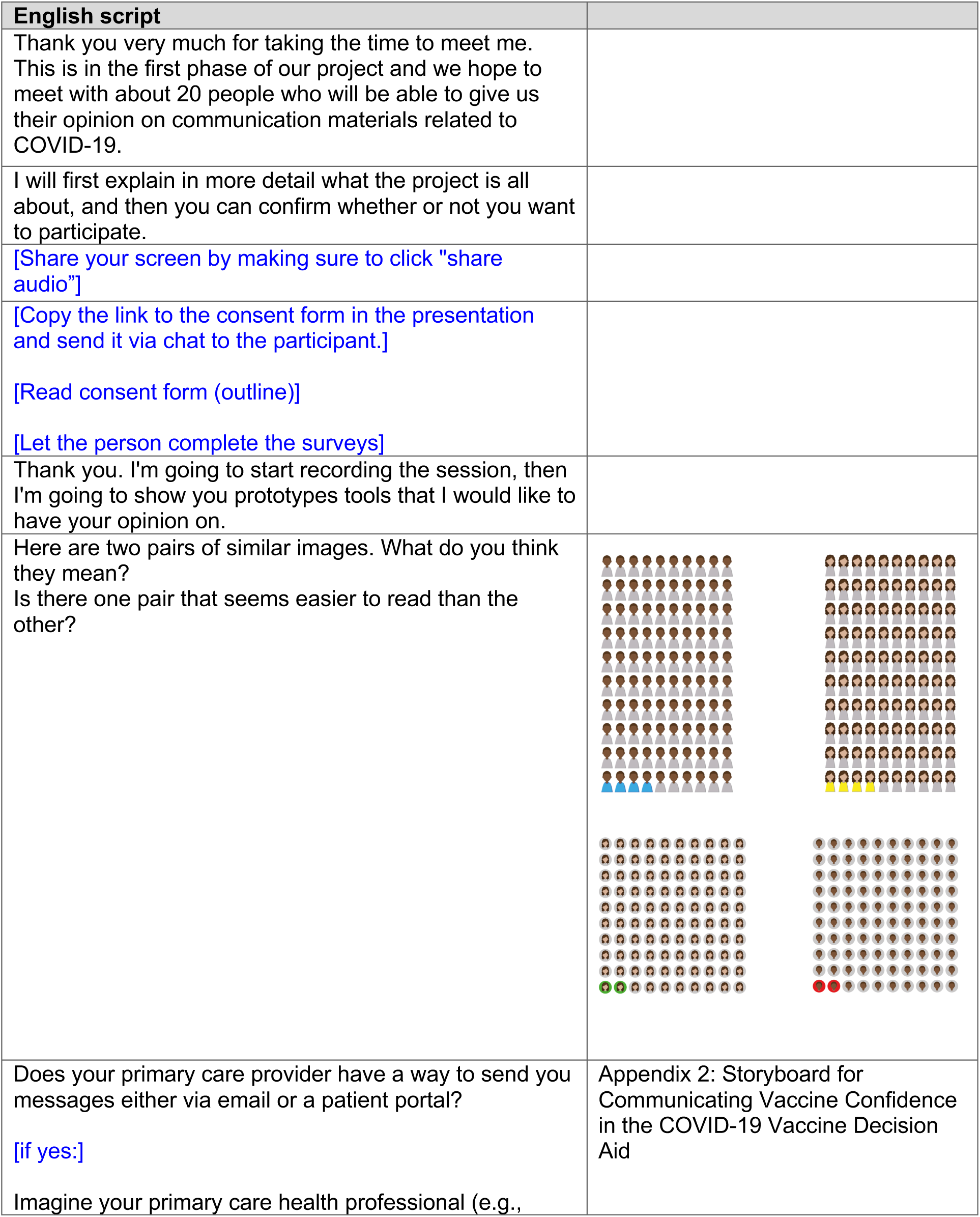
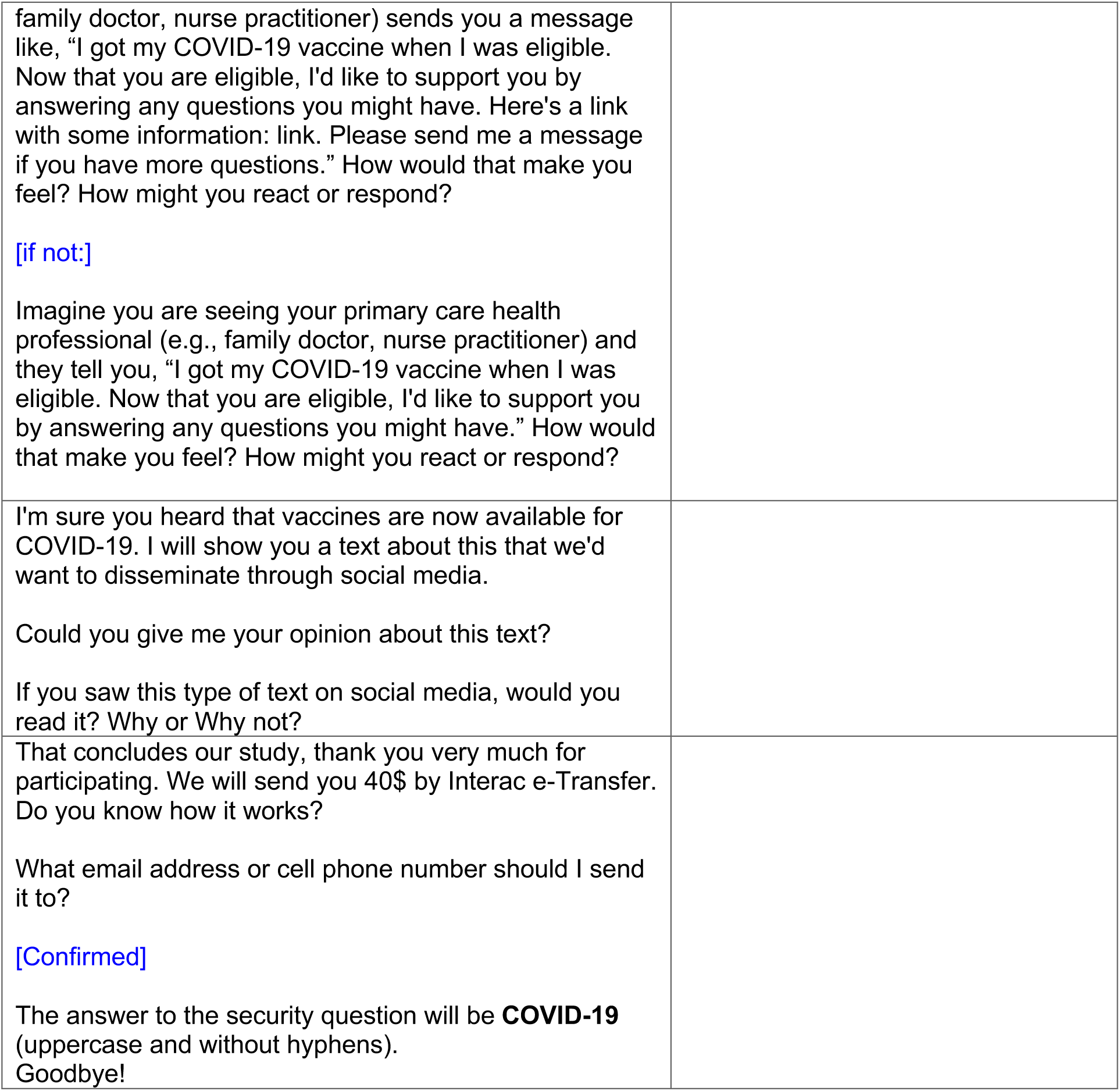

| English script |  |
| --- | --- |
| Thank you very much for taking the time to meet me. This is in the first phase of our project and we hope to meet with about 20 people who will be able to give us their opinion on communication materials related to COVID-19. |  |
| I will first explain in more detail what the project is all about, and then you can confirm whether or not you want to participate. |  |
| [Share your screen by making sure to click "share audio"] |  |
| [Copy the link to the consent form in the presentation and send it via chat to the participant.]<br>[Read consent form (outline)]<br>[Let the person complete the surveys] |  |
| Thank you. I'm going to start recording the session, then I'm going to show you prototypes tools that I would like to have your opinion on. |  |
| Here are two pairs of similar images. What do you think they mean?<br>Is there one pair that seems easier to read than the other? |  |
| Does your primary care provider have a way to send you messages either via email or a patient portal?<br>[if yes:]<br>Imagine your primary care health professional (e.g., | Appendix 2: Storyboard for Communicating Vaccine Confidence in the COVID-19 Vaccine Decision Aid |
| family doctor, nurse practitioner) sends you a message like, "I got my COVID-19 vaccine when I was eligible. Now that you are eligible, I'd like to support you by answering any questions you might have. Here's a link with some information: link. Please send me a message if you have more questions." How would that make you feel? How might you react or respond?<br>[if not:]<br>Imagine you are seeing your primary care health professional (e.g., family doctor, nurse practitioner) and they tell you, "I got my COVID-19 vaccine when I was eligible. Now that you are eligible, I'd like to support you by answering any questions you might have." How would that make you feel? How might you react or respond? |  |
| I'm sure you heard that vaccines are now available for COVID-19. I will show you a text about this that we'd want to disseminate through social media.<br>Could you give me your opinion about this text?<br>If you saw this type of text on social media, would you read it? Why or Why not? |  |
| That concludes our study, thank you very much for participating. We will send you 40\$ by Interac e-Transfer. Do you know how it works?<br>What email address or cell phone number should I send it to?<br>[Confirmed]<br>The answer to the security question will be **COVID-19** (uppercase and without hyphens).<br>Goodbye! | |

### Appendix 2. Proposed social media text on vaccine confidence - Cycle 1

Text and image description for an intervention that will be produced as both a series of images (for use on platforms where multiple images work best, e.g., tweet thread, Instagram carousel) or an infographic (e.g., Facebook, Pinterest)

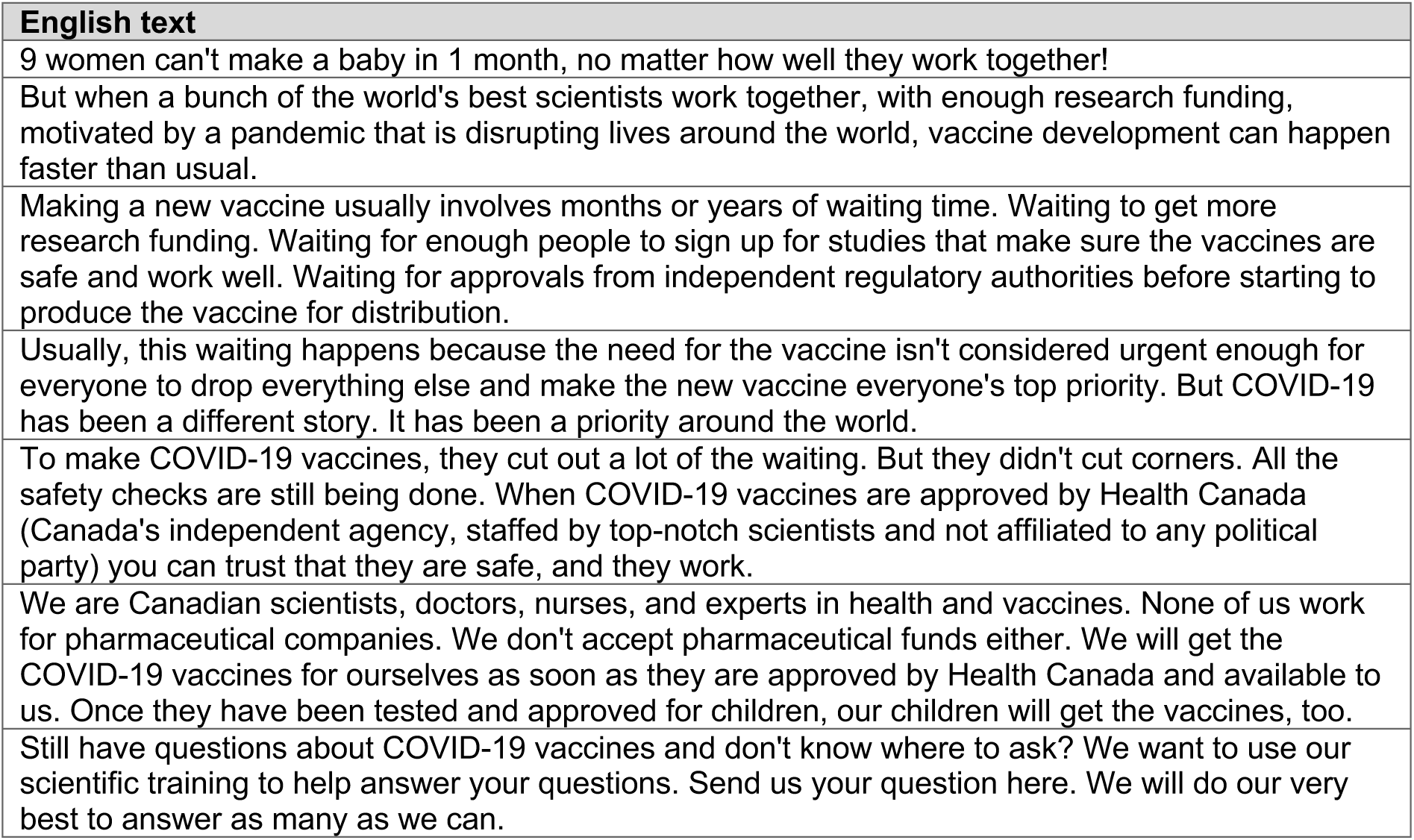

### Appendix 3. User testing interview guide - Cycle 2

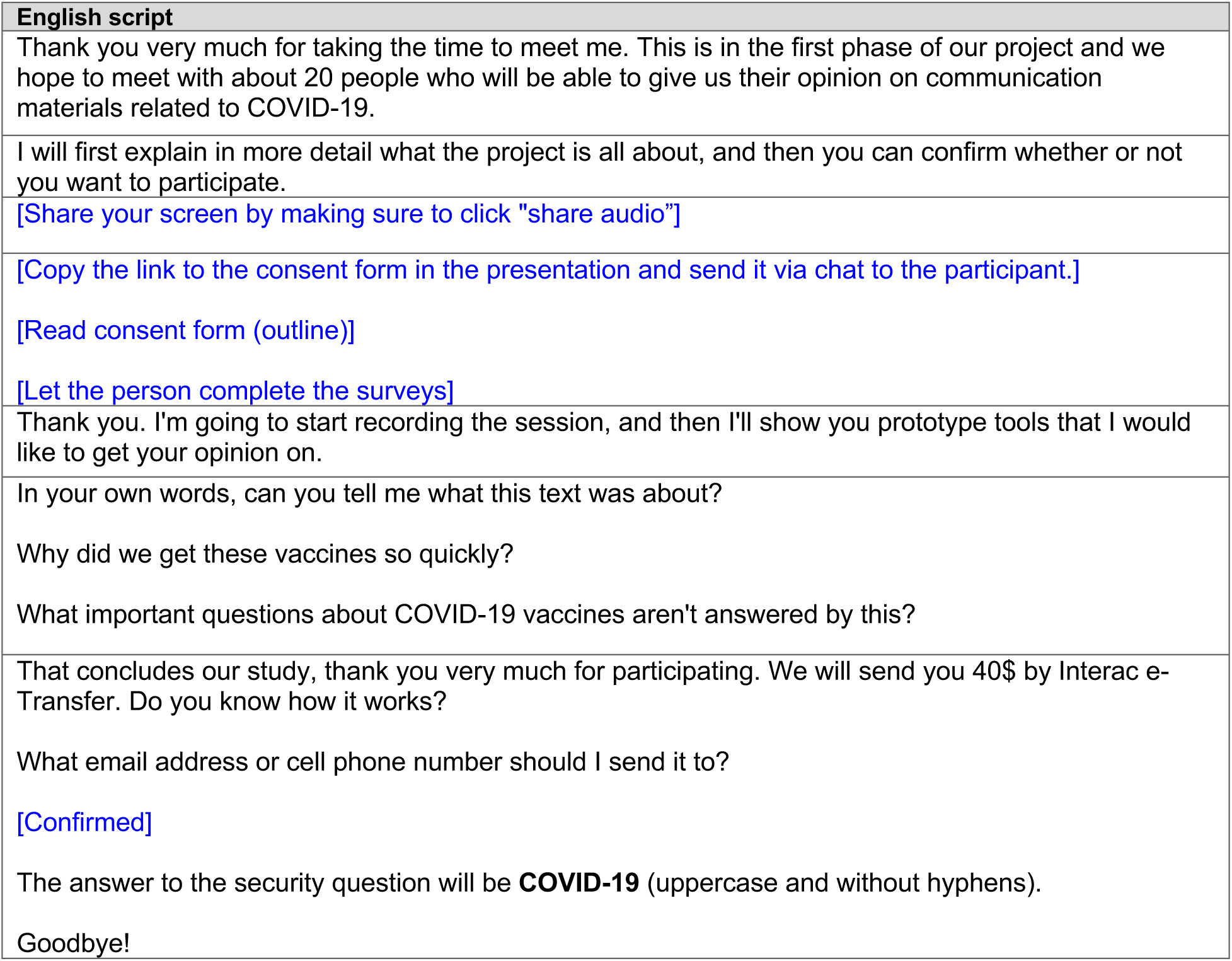

| English script |
| --- |
| Thank you very much for taking the time to meet me. This is in the first phase of our project and we hope to meet with about 20 people who will be able to give us their opinion on communication materials related to COVID-19. |
| I will first explain in more detail what the project is all about, and then you can confirm whether or not you want to participate. |
| [Share your screen by making sure to click "share audio"] |
| [Copy the link to the consent form in the presentation and send it via chat to the participant.] |
| [Read consent form (outline)] |
| [Let the person complete the surveys] |
| Thank you. I'm going to start recording the session, and then I'll show you prototype tools that I would like to get your opinion on. |
| In your own words, can you tell me what this text was about? |
| Why did we get these vaccines so quickly? |
| What important questions about COVID-19 vaccines aren't answered by this? |
| That concludes our study, thank you very much for participating. We will send you 40\$ by Interac e-Transfer. Do you know how it works? |
| What email address or cell phone number should I send it to? |
| [Confirmed] |
| The answer to the security question will be **COVID-19** (uppercase and without hyphens). |
| Goodbye! |

### Appendix 4. Vaccine explanation text storyboard used in Cycle 2

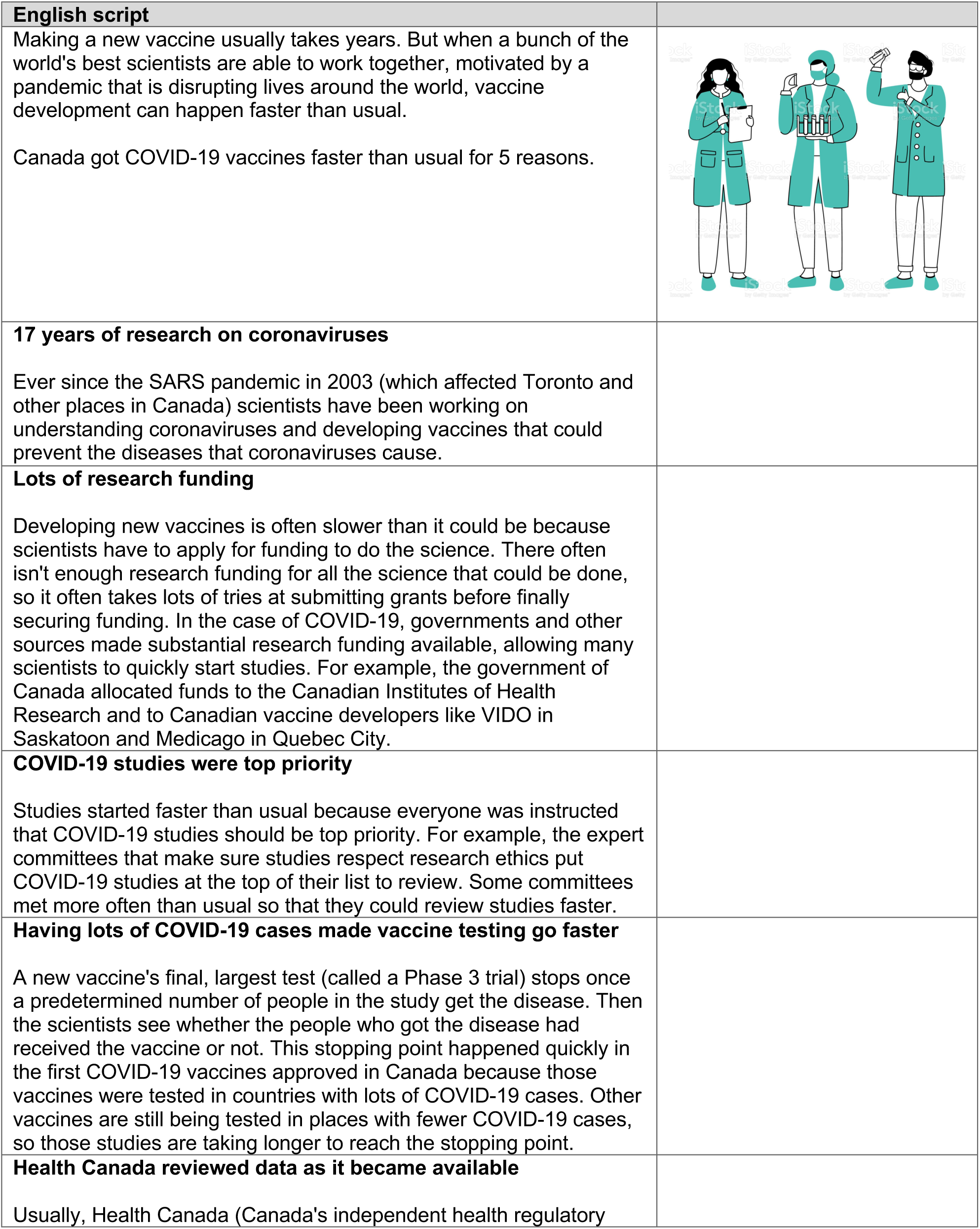
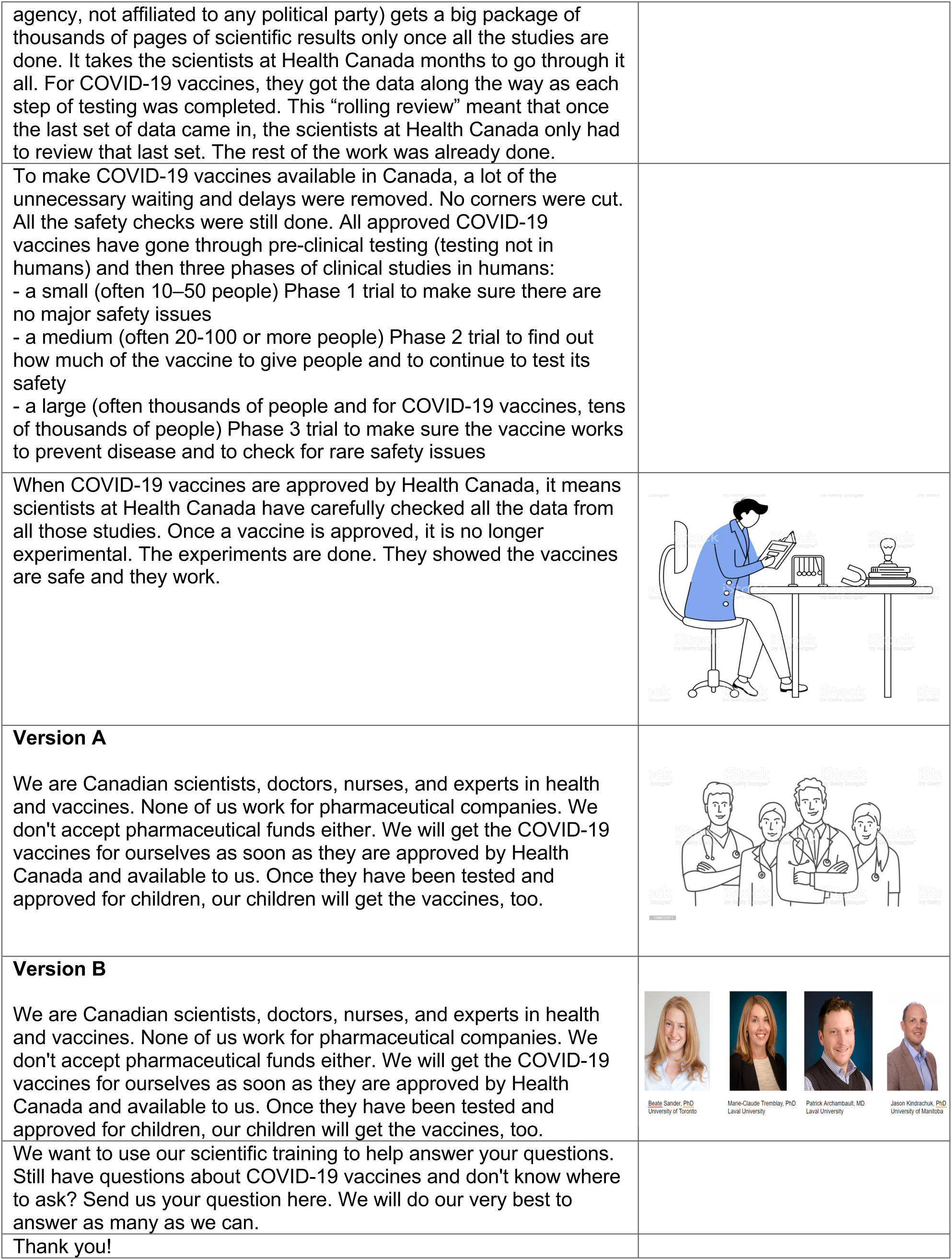

|  |
| --- |
| **English script**<br>Making a new vaccine usually takes years. But when a bunch of the world's best scientists are able to work together, motivated by a pandemic that is disrupting lives around the world, vaccine development can happen faster than usual.<br>Canada got COVID-19 vaccines faster than usual for 5 reasons. |
| **17 years of research on coronaviruses**<br>Ever since the SARS pandemic in 2003 (which affected Toronto and other places in Canada) scientists have been working on understanding coronaviruses and developing vaccines that could prevent the diseases that coronaviruses cause. |
| **Lots of research funding**<br>Developing new vaccines is often slower than it could be because scientists have to apply for funding to do the science. There often isn't enough research funding for all the science that could be done, so it often takes lots of tries at submitting grants before finally securing funding. In the case of COVID-19, governments and other sources made substantial research funding available, allowing many scientists to quickly start studies. For example, the government of Canada allocated funds to the Canadian Institutes of Health Research and to Canadian vaccine developers like VIDO in Saskatoon and Medicago in Quebec City. |
| **COVID-19 studies were top priority**<br>Studies started faster than usual because everyone was instructed that COVID-19 studies should be top priority. For example, the expert committees that make sure studies respect research ethics put COVID-19 studies at the top of their list to review. Some committees met more often than usual so that they could review studies faster. |
| **Having lots of COVID-19 cases made vaccine testing go faster**<br>A new vaccine's final, largest test (called a Phase 3 trial) stops once a predetermined number of people in the study get the disease. Then the scientists see whether the people who got the disease had received the vaccine or not. This stopping point happened quickly in the first COVID-19 vaccines approved in Canada because those vaccines were tested in countries with lots of COVID-19 cases. Other vaccines are still being tested in places with fewer COVID-19 cases, so those studies are taking longer to reach the stopping point. |
| **Health Canada reviewed data as it became available**<br>Usually, Health Canada (Canada's independent health regulatory |

|  |  |
| --- | --- |
| agency, not affiliated to any political party) gets a big package of thousands of pages of scientific results only once all the studies are done. It takes the scientists at Health Canada months to go through it all. For COVID-19 vaccines, they got the data along the way as each step of testing was completed. This “rolling review” meant that once the last set of data came in, the scientists at Health Canada only had to review that last set. The rest of the work was already done. |  |
| To make COVID-19 vaccines available in Canada, a lot of the unnecessary waiting and delays were removed. No corners were cut. All the safety checks were still done. All approved COVID-19 vaccines have gone through pre-clinical testing (testing not in humans) and then three phases of clinical studies in humans:<br>- a small (often 10–50 people) Phase 1 trial to make sure there are no major safety issues<br>- a medium (often 20-100 or more people) Phase 2 trial to find out how much of the vaccine to give people and to continue to test its safety<br>- a large (often thousands of people and for COVID-19 vaccines, tens of thousands of people) Phase 3 trial to make sure the vaccine works to prevent disease and to check for rare safety issues |  |
| When COVID-19 vaccines are approved by Health Canada, it means scientists at Health Canada have carefully checked all the data from all those studies. Once a vaccine is approved, it is no longer experimental. The experiments are done. They showed the vaccines are safe and they work. |  |
| **Version A**<br>We are Canadian scientists, doctors, nurses, and experts in health and vaccines. None of us work for pharmaceutical companies. We don't accept pharmaceutical funds either. We will get the COVID-19 vaccines for ourselves as soon as they are approved by Health Canada and available to us. Once they have been tested and approved for children, our children will get the vaccines, too. |  |
| **Version B**<br>We are Canadian scientists, doctors, nurses, and experts in health and vaccines. None of us work for pharmaceutical companies. We don't accept pharmaceutical funds either. We will get the COVID-19 vaccines for ourselves as soon as they are approved by Health Canada and available to us. Once they have been tested and approved for children, our children will get the vaccines, too. | Beate Sander, PhD<br>University of Toronto<br>Marie-Claude Tremblay, PhD<br>Laval University<br>Patrick Archambault, MD<br>Laval University<br>Jason Kindrachuk, PhD<br>University of Manitoba |
| We want to use our scientific training to help answer your questions. Still have questions about COVID-19 vaccines and don't know where to ask? Send us your question here. We will do our very best to answer as many as we can. |  |
| Thank you! |  |

### Appendix 5. Technical development and version control workflow for VaxDA-C19

Our development team used a structured Git workflow to manage and document the iterative build of VaxDA-C19. We organized the codebase into three main branches: the Dev Branch (249 commits) for implementing new features and experimental changes; the NewDesign Branch (367 commits) for redesigning the user interface based on usability testing feedback; and the Master Branch (308 commits) for stable, production-ready versions. This structure allowed the team to implement updates rapidly, track changes precisely, and maintain proper version histories throughout the project.

We integrated user feedback from each testing cycle directly into the development stream. Developers tagged and documented all modifications related to visual layout, navigation logic, content rendering, and internalization. We relied on JSON-based templates to allow dynamic content updates and easy localization in both English and French.

To ensure reproducibility and transparency, the team mirrored the staging and production environments and followed continuous integration practices to detect bugs early and maintain deployment stability. This technical framework allowed us to integrate public health guidance changes while preserving the accuracy and consistency of the patient decision aid.

## Abbreviations

CAD: Canadian dollar
COVID-19: coronavirus disease 2019
CSS: Cascading Style Sheets
EPPM: Extended Parallel Process Model
FAQs: Frequently Asked Questions
HTML: Hyper Text Markup Language
JSON: JavaScript Object Notation
PR: Public relations
SMDM: Society for Medical Decision-Making
US: United States
VaxDA-C19: web-based vaccine (Vax) patient decision aid (DA) designed to support COVID-19 (-C19) vaccination

## DECLARATIONS

### Availability of Data and Materials

All data generated or analyzed during this study are included in this published article and are available on demand. The French versions of Appendices 1–3 are not included per medRxiv policy. They are available from the corresponding author upon reasonable request.

## Authors’ Contributions

DE, HW, and ED contributed to the design of the study. DE, EP, HH and HW contributed to data collection. DE conducted data analysis and interpretation. DE and HW drafted the first version of the article with early revision. PA, IB, CTC, ADC, JD, SMD, ED, MPG, TG, AG, NG, KG, HH, SJ, JK, AL, SEM, RN, MN, RO, JSP, EP, JSR, BS, MT, DTG, MCT, SVM, VW critically revised the article and approved the final version for submission for publication. DE and HW had full access to all the data in the study and had final responsibility for the decision to submit for publication.

## Acknowledgements

The authors gratefully acknowledge the contributions of all the team members who contributed greatly to this research project as a whole but were unable to accept the invitation to participate on this particular paper as co-authors due to time constraints, including citizen panel members who chose not to accept the invitation to be co-authors: Ross McCreery, Samantha Meyer, Linda Kim, Jean Légaré, and Melissa McLeod. We thank Magniol Noubi and Dan Emmanuel Marie Krecoum for programming the web application and Selma Chipenda Dansokho for constructive comments throughout the project. Finally, we thank all study participants for helping us identify ways to improve education, communication, and decision-making in health care.

